# Comprehensive lipidomics reveals reduced hepatic lipid turnover in NAFLD during alcohol intoxication

**DOI:** 10.1101/2021.02.18.21251910

**Authors:** Mads Israelsen, Min Kim, Tommi Suvitaival, Bjørn Stæhr Madsen, Camilla Dalby Hansen, Nikolaj Torp, Kajetan Trost, Maja Thiele, Torben Hansen, Cristina Legido-Quigley, Aleksander Krag, on behalf of the MicrobLiver Consortium

## Abstract

**Background & Aims:** In experimental models, alcohol induces acute changes in lipid metabolism that cause hepatocyte lipoapoptosis and inflammation. Here we study human hepatic lipid turnover during controlled alcohol intoxication.

**Methods:** We studied 39 participants with three distinct hepatic phenotypes: alcohol-related liver disease (ALD), non-alcoholic fatty liver disease (NAFLD), and healthy controls. Alcohol was administrated via nasogastric tube over 30 minutes. Hepatic and systemic venous blood were sampled simultaneously at three time points: baseline, 60 and 180 min after alcohol intervention. Liver biopsies were sampled 240 minutes after alcohol intervention. We used ultra-high-performance liquid chromatograph mass spectrometry to measure levels of more than 250 lipid species from the blood and liver samples.

**Results:** After alcohol intervention, the levels of blood free fatty acid (FFA) and lysophosphatidylcholine (LPC) decreased while triglyceride (TG) increased. FFA was the only lipid class to decrease in NAFLD after alcohol intervention, while LPC and FFA decreased and TG increased after intervention in ALD and healthy controls. Fatty acid chain uptake preference in FFAs and LPCs were oleic acid, linoleic acid, arachidonic acid, and docosahexaenoic acid. Hepatic venous blood FFA and LPC levels were lower when compared to systemic venous blood levels throughout the intervention. After alcohol intoxication, liver lipidome in ALD was similar to that in NAFLD.

**Conclusions:** Alcohol intoxication induces rapid changes in circulating lipids including hepatic turnaround from FFA and LPC, potentially leading to lipoapoptosis and steatohepatitis. TG clearance was suppressed in NAFLD, possibly explaining why alcohol and NAFLD are synergistic risk factors for disease progression. These effects may be central to the pathogenesis of ALD.

## Introduction

Alcohol is the leading etiology of cirrhosis and is associated with high morbidity and mortality.^1^ From 1999 to 2016 the United States experienced a 2-10% annual increase in mortality due to alcohol-related liver disease (ALD).^2^ Worldwide 20% of all people have at least one heavy drinking episode per month^3^ and weakly heavy drinking episodes are seen in 13% of people in the United States.^4^ Heavy drinking episodes are associated with higher risk of developing chronic liver injury and alcohol-related cirrhosis,^5^ with a particular high risk in obese people.^6-8^

Alcohol-induced lipo-toxicity is an emerging area and have an important role in the pathogenesis of alcohol-related steatohepatitis. Hepatic alcohol detoxification leads to formation of toxic metabolites and hepatic steatosis, and it is well-established that alcohol causes hepatic accumulation of FFAs by blocking mitochondrial beta-oxidation.^9^ However, experimental models suggest a more complex interaction between alcohol and hepatic lipid metabolism; alcohol increases the hepatic uptake of FFA, alcohol has different effects on distinct types of FFA (short versus long-chain and saturated versus unsaturated), and alcohol may also affect other lipid classes including lysophosphatidylcholines (LPCs) that can induce lipo-toxicity.^9, 10^ Furthermore, animal models reveal that high fat diet increases the susceptibility of alcohol-induced injury and the combination leads to increased progression rate of chronic liver injury.^11^

Although experimental studies have revealed mechanistic knowledge, we lack human studies that can aid the clinical understanding of lipid metabolism in relation to alcohol intoxication. To investigate the translation of experimental findings to humans, we developed a study where participants were given a controlled amount of alcohol to replicate alcohol intoxication (Figure 1A). We recruited participants with three different hepatic phenotypes: healthy control, ALD, and non-alcoholic fatty liver disease (NAFLD). Blood samples from hepatic and systemic veins were collected before and after alcohol intoxication from all three hepatic phenotypes. Liver biopsy samples were collected by the end of alcohol intoxication from participants with ALD and NAFLD. Comprehensive lipidomic analyzes were performed on these blood and liver tissue samples (Figure 1B). Our overall aims were to (1) explore the effects of alcohol intoxication on the hepatic venous blood lipid profile, (2) test whether alcohol intoxication causes different reactions in people with ALD or NAFLD and healthy controls, and (3) explore differences between hepatic and systemic blood lipid profiles as well as liver lipid profiles.

**Figure 1.**
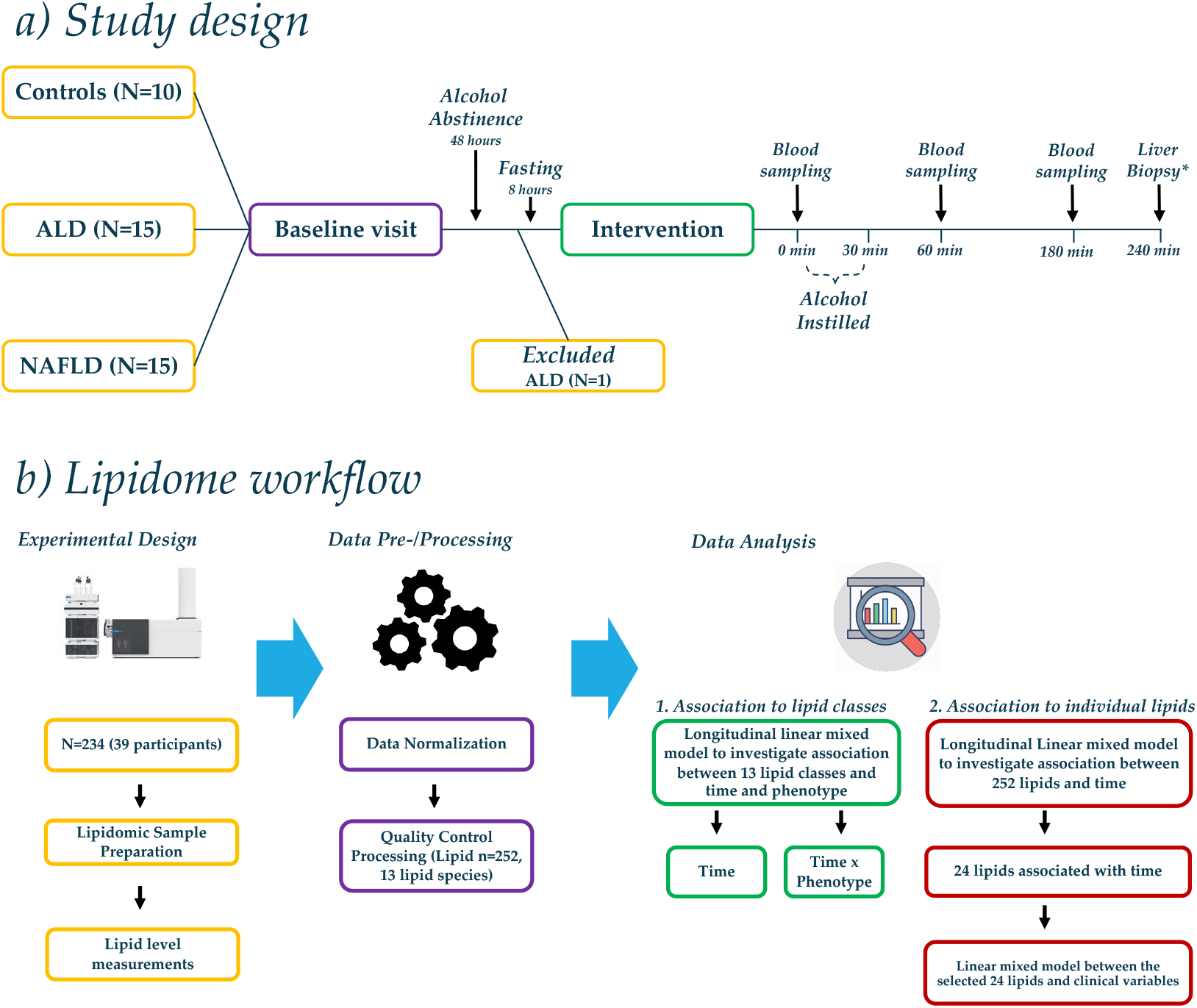
Schematic study design and Lipidome workflow. **A) Study design**: 40 individuals came for baseline visit, but one ALD participant was subsequently excluded due to protocol violation by consuming alcohol within 48 hours prior to the alcohol intervention. The 39 participants had three distinct hepatic phenotypes: 10 healthy controls, 14 with ALD, and 15 with NAFLD. Blood was sampled at time 0 minutes, and alcohol was subsequently instilled over 30 minutes. Blood was sampled again after 60 and 180 minutes. *Transjugular liver biopsies were collected after 240 minutes in participants with ALD and NAFLD **B) Lipidome workflow:** Lipid levels (n=252) were measured using ultra-high-performance liquid chromatograph coupled with quadrupole time-of-flight mass spectrometer (UHPLC-QTOFMS). First, we explored the average level of 13 distinct lipid classes from hepatic venous blood to identify which lipid class changed in its level over time after alcohol intervention. We used longitudinal mixed effect models to investigate three different fixed effects: (1) ‘time’, (2) ‘time*phenotype’, and (3) ‘time*blood site’. Fixed effect of ‘time’ allowed us to identify lipid classes that changed in their levels after alcohol intervention. Fixed effect of ‘time*phenotype’ allowed us to see whether levels of 13 lipid classes were different between the hepatic phenotypes before and after alcohol intervention. Fixed effect of ‘time*blood site’ allowed us to see whether levels of 13 lipid classes were different between two blood vein sites (systemic vs. hepatic) before and after alcohol intervention. Finally, we investigated 252 individual lipid species using the same approach where fixed effects were (1) ‘time’, (2) ‘time*phenotype’, and (3) ‘time*blood site’. In all models, random effect was the individual participant.

## Results

### Participants

Baseline investigations were performed on 40 individuals from November 2016 to November 2018, but one participant with ALD was subsequently excluded due to protocol violation by consuming alcohol within 48 hours of the alcohol intervention. Therefore, 39 participants with three distinct hepatic phenotypes were included: ALD (n=14), NAFLD (n=15), and healthy controls (n=10). Table 1 shows the participants’ baseline characteristics. The mean age was 54 ±11 years, and 62% were males, and there were no significant differences between the groups. Median alcohol consumption was 60 g/day in ALD and below 12 g/day in NAFLD and healthy controls (p<0.001). NAFLD participants had higher BMI, higher HbA1C, and lower HDL cholesterol than ALD and healthy controls (p<0.01). NAFLD participants also had significantly higher insulin and HOMA-IR than healthy controls. There was no difference between ALD and NAFLD with regards to liver blood tests and liver histopathology (Table 1). Overall, 55% had significant fibrosis (≥F2), 48% had moderate or severe steatosis (≥S2), and the median NAFLD activity score was 3. All the healthy controls had normal liver stiffness (<6 kPa).

**Table 1:**
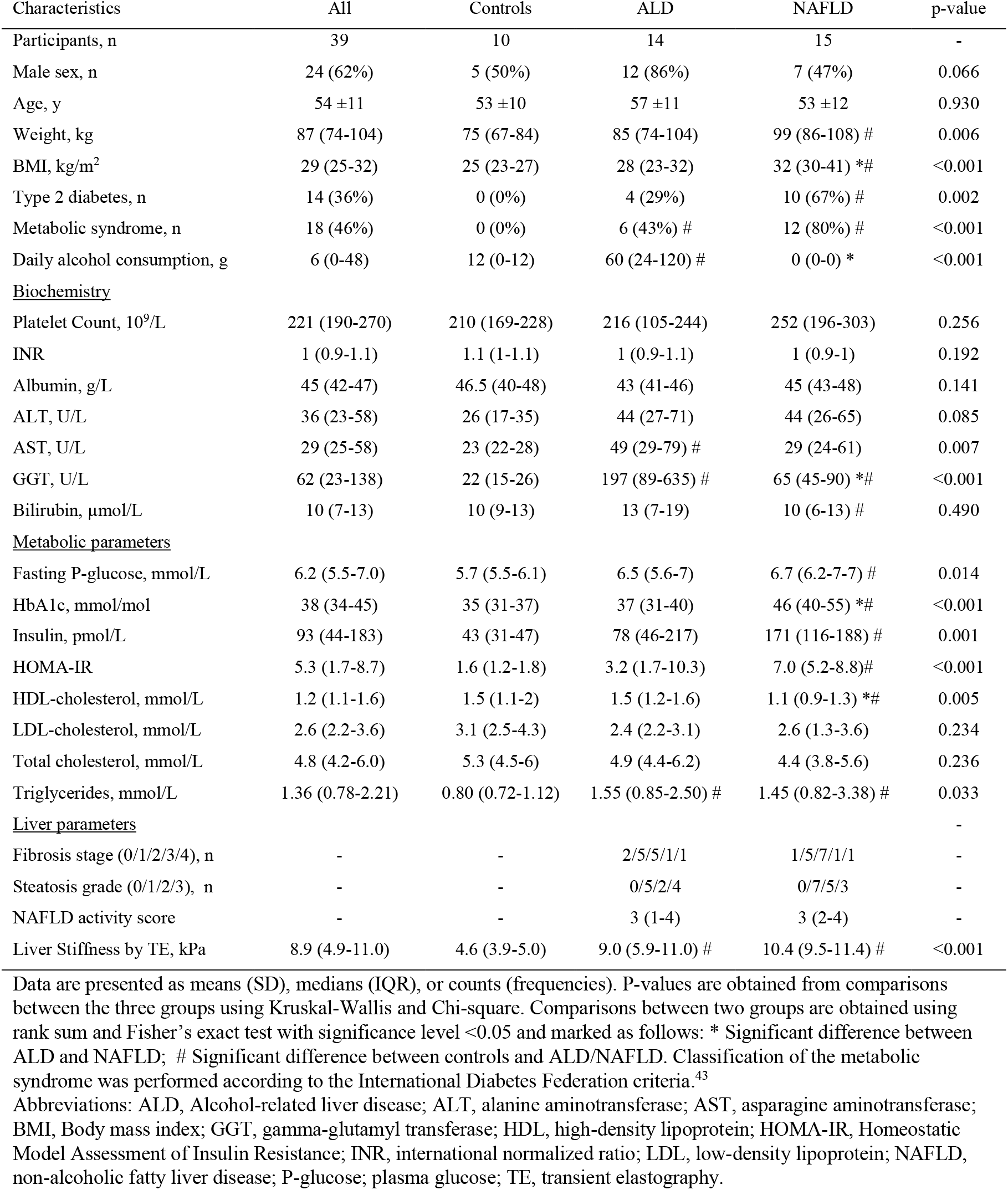
Participant characteristics.

### Alcohol intervention

All participants received the scheduled alcohol dose according to protocol. The mean serum alcohol concentration was 0±0 mmol/L (0.00±0.0 g/dL) at baseline, peaked at 34±4 mmol/L (0.16±0.02 g/dL) after 60 min, and declined to 21±3 (0.10±0.1 g/dL) mmol/L after 180 min (p<0.001). Only mild transient symptoms commonly seen in relation to alcohol intake were observed.

### Hepatic venous lipid classes before and after alcohol intervention

Before alcohol intervention, healthy controls had lower hepatic venous triglyceride (TG) levels compared to ALD and NAFLD (Figure 2a and Table 3). Alcohol intervention led to significant reductions in levels of FFA (−39.42 units per minute, p<0.0001) and LPC (−14.08 units per minute, p=0.0002) (Table 2 and Figure 2A), but not for other lipid classes. Supplementary Table 1 summarizes the effect of alcohol intervention on all 13 lipid classes.

**Table 2.**
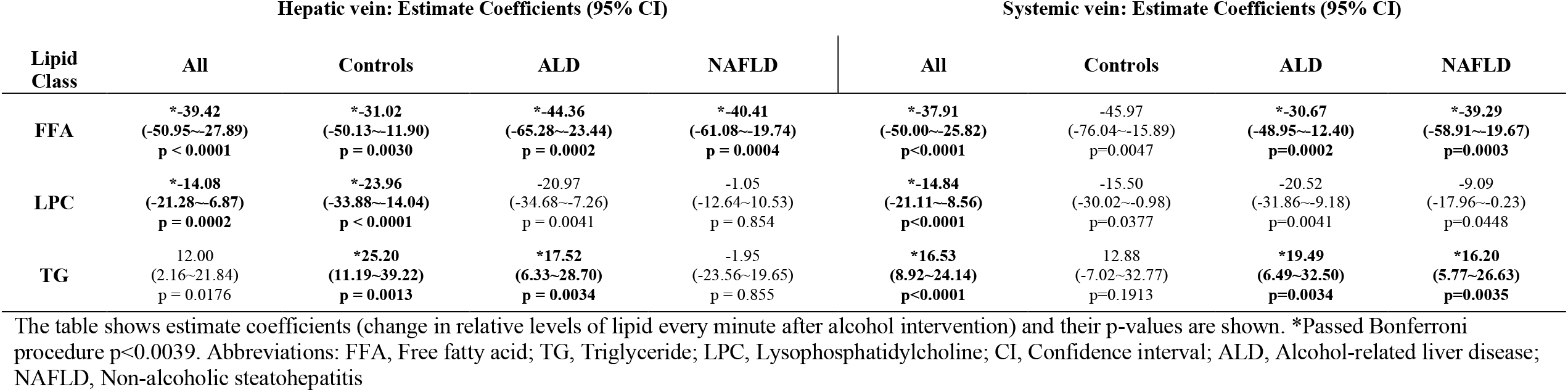
Summary of mixed models showing relative changes in FFA, LPC, and TG levels from baseline to 180 min after start of alcohol intake.

| Lipid Class | Hepatic vein: Estimate Coefficients (95% CI) |  |  |  | Systemic vein: Estimate Coefficients (95% CI) |  |  |  |
| --- | --- | --- | --- | --- | --- | --- | --- | --- |
|  | All | Controls | ALD | NAFLD | All | Controls | ALD | NAFLD |
| **FFA** | **\*-39.42**<br>(-50.95~-27.89)<br>**p < 0.0001** | **\*-31.02**<br>(-50.13~-11.90)<br>**p = 0.0030** | **\*-44.36**<br>(-65.28~-23.44)<br>**p = 0.0002** | **\*-40.41**<br>(-61.08~-19.74)<br>**p = 0.0004** | **\*-37.91**<br>(-50.00~-25.82)<br>**p<0.0001** | -45.97<br>(-76.04~-15.89)<br>p=0.0047 | **\*-30.67**<br>(-48.95~-12.40)<br>**p=0.0002** | **\*-39.29**<br>(-58.91~-19.67)<br>**p=0.0003** |
| **LPC** | **\*-14.08**<br>(-21.28~-6.87)<br>**p = 0.0002** | **\*-23.96**<br>(-33.88~-14.04)<br>**p < 0.0001** | -20.97<br>(-34.68~-7.26)<br>p = 0.0041 | -1.05<br>(-12.64~10.53)<br>p = 0.854 | **\*-14.84**<br>(-21.11~-8.56)<br>**p<0.0001** | -15.50<br>(-30.02~-0.98)<br>p=0.0377 | -20.52<br>(-31.86~-9.18)<br>p=0.0041 | -9.09<br>(-17.96~-0.23)<br>p=0.0448 |
| **TG** | 12.00<br>(2.16~21.84)<br>p = 0.0176 | **\*25.20**<br>(11.19~39.22)<br>**p = 0.0013** | **\*17.52**<br>(6.33~28.70)<br>**p = 0.0034** | -1.95<br>(-23.56~19.65)<br>p = 0.855 | **\*16.53**<br>(8.92~24.14)<br>**p<0.0001** | 12.88<br>(-7.02~32.77)<br>p=0.1913 | **\*19.49**<br>(6.49~32.50)<br>**p=0.0034** | **\*16.20**<br>(5.77~26.63)<br>**p=0.0035** |

The table shows estimate coefficients (change in relative levels of lipid every minute after alcohol intervention) and their p-values are shown. \*Passed Bonferroni procedure $p < 0.0039$ . Abbreviations: FFA, Free fatty acid; TG, Triglyceride; LPC, Lysophosphatidylcholine; CI, Confidence interval; ALD, Alcohol-related liver disease; NAFLD, Non-alcoholic steatohepatitis

**Table 3.**
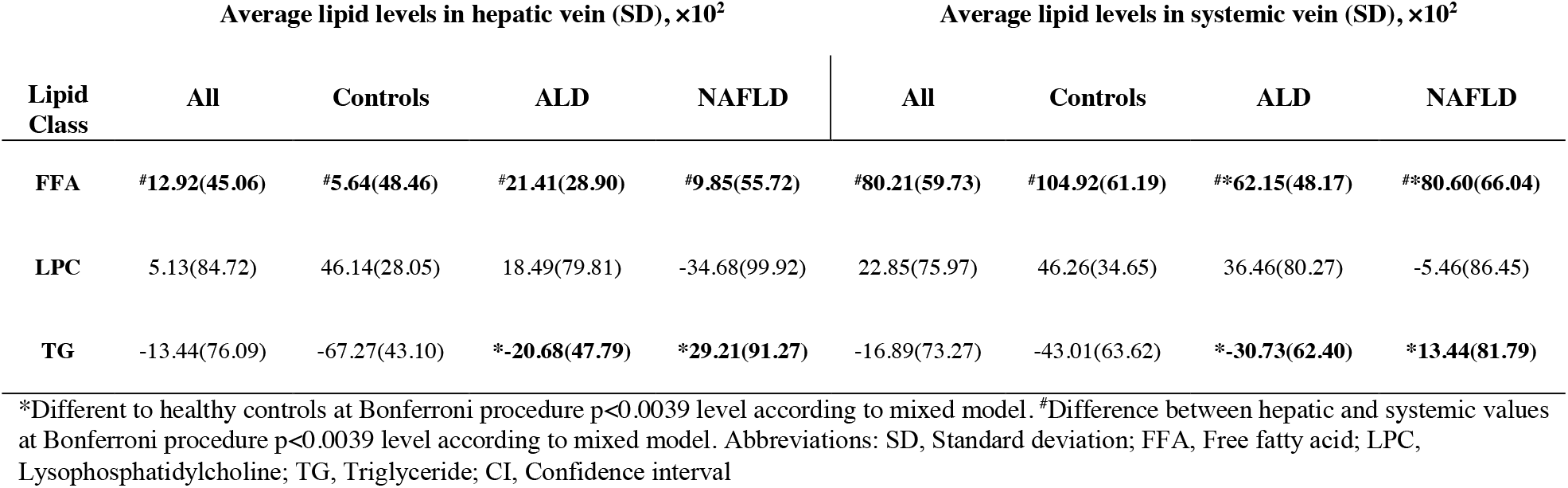
Comparison of FFA, LPC and TG at baseline between hepatic and systemic venous blood.

**Figure 2.**
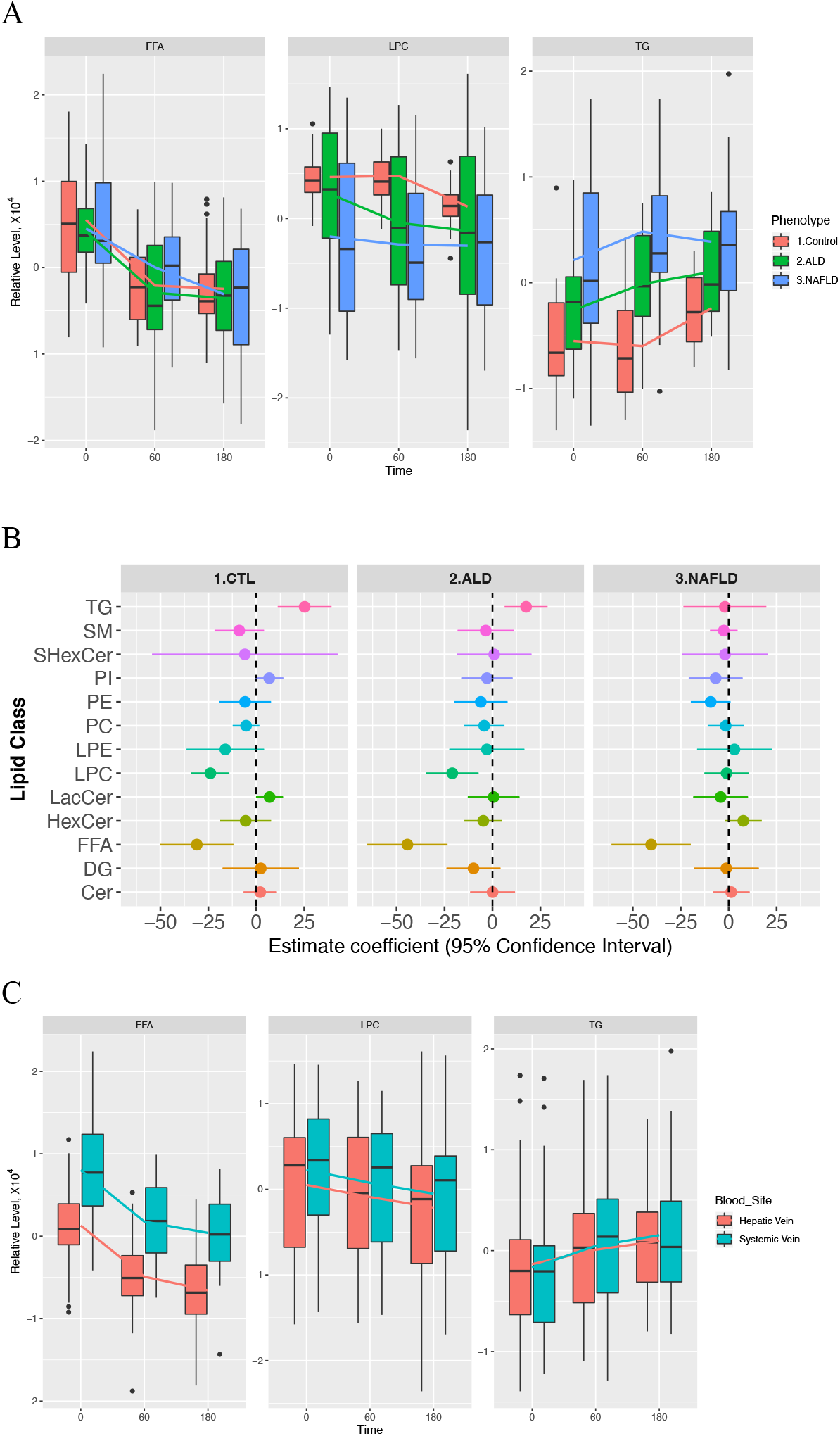
(A) Boxplots showing shows FFA, LPC and TG levels from hepatic vein for the three hepatic phenotypes. These three lipid classes were selected here because their levels changed after alcohol intervention in at least one phenotype (Table 2). FFA showed significant changes over time in Control, ALD, and NAFLD phenotypes, while LPC showed significant changes over time in Control phenotype. TG showed significant changes over time in ALD and NAFLD phenotypes. Significances were based on p<0.0039. (B) Forrest plot showing magnitude of change after alcohol intervention (estimate coefficients) for all 13 lipid classes from hepatic vein in each hepatic phenotype. (C) Boxplots comparing FFA, LPC, and TG levels in hepatic and systemic veins. The lines connect the means of boxes.

### Effect of alcohol intervention by phenotype on hepatic venous lipid classes

Alcohol caused significant reductions in hepatic venous FFA levels in all three hepatic phenotypes (Figure 2B): healthy controls (−31.02 units per minute, p=0.0030), ALD (−44.36 units per minute, p=0.0002), and NAFLD (−40.41 units per minute, p=0.0004) (Figure 2B and Table 2). In contrast, LPC decreased only in healthy controls (−23.96 units per minute, p<0.0001) and in ALD (−20.97 units per minute, p=0.0041) (Table 2). Hepatic TG levels increased significantly in healthy controls (+25.20 units per minute, p=0.0013) and in ALD (+17.52 units per minute, p=0.0034) but not in NAFLD (−1.95 units per minute, p=0.855) (Figure 2B and Table 2).

Estimate coefficient values for each lipid class and between-group comparisons are provided in Supplementary Tables 1 and 2.

### Hepatic versus systemic venous blood

Before alcohol intervention, the level of FFA in all three hepatic phenotypes was significantly lower in hepatic venous blood than in systemic venous blood (Figure 2C and Table 3). However, the rates at which lipid classes changed during alcohol intervention (estimate coefficient values) did not differ between hepatic and systemic venous blood for any of the lipid classes (Supplementary Table 1).

### Single lipid levels before and after alcohol intervention

Twenty-four of 252 individual lipids changed significantly in hepatic venous blood after alcohol intervention at Bonferroni level of p<1.98×10^−4^ (Figure 3A). Estimate coefficient values for these 24 lipids can be found in Supplementary Table 4. One mono-unsaturated fatty acid (MUFA) and three poly-unsaturated fatty acids (PUFAs) decreased in response to alcohol: oleic acid (FFA 18:1), linoleic acid (FFA 18:2), arachidonic acid (FFA 20:4), and docosahexaenoic acid (FFA 22:6). The levels of nine LPCs decreased, four of which contained the same four FFAs as above. The nine LPC molecules contained oleic acid (FFA 18:1), linoleic acid (FFA 18:2), arachidonic acid (FFA 20:4), dihomo-α-linolenic (FFA 20:3), docosapentaenoic acid (FFA 20:5), docosapentaenoic acid (FFA 22:5), and docosahexaenoic acid (FFA 22:6) (Figure 3B). The alcohol intervention also led to increased levels of one phosphatidylcholine (PC) and 10 TG molecules.

**Figure 3.**
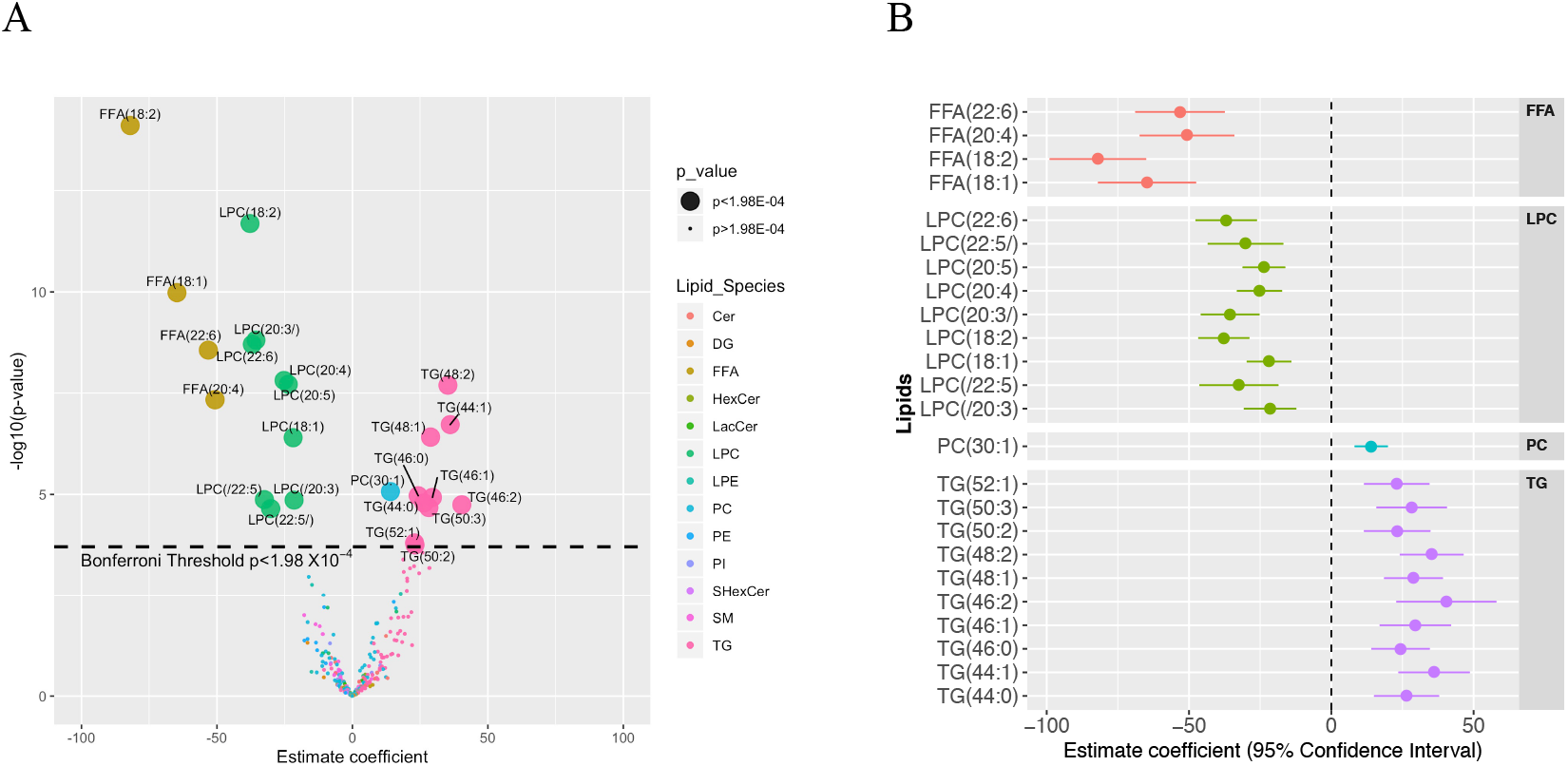
(A) Volcano plot showing changes in levels (estimate coefficients) for 252 lipids from hepatic vein after alcohol intervention. Statistical significance (y-axis) is plotted against the magnitude of change in the lipid levels after alcohol intervention (x-axis) to identify the most dynamic lipids that were also statistically significant. Different colours represent different lipid species, and larger dots represent the 24 lipids passing Bonferroni threshold (p<1.98×10^−04^). (B) Forrest plot showing magnitude of change after alcohol intervention (estimate coefficients) for the 24 lipids from hepatic vein selected from Figure 3A.

### Single lipid levels by phenotype before and after alcohol intervention

We focused on these 24 lipids and compared their levels between the different hepatic phenotype groups at baseline. The healthy controls had significantly lower levels of all 10 TG lipids compared to ALD while seven TG lipids were also significantly lower in healthy controls compared to NAFLD (Figure 4 and Supplementary Table 4). No lipids differed between ALD and NAFLD before alcohol intervention (Supplementary Table 5). The four FFA levels were similar in healthy controls, ALD, and NAFLD before alcohol intervention (Figure 4).

**Figure 4.**
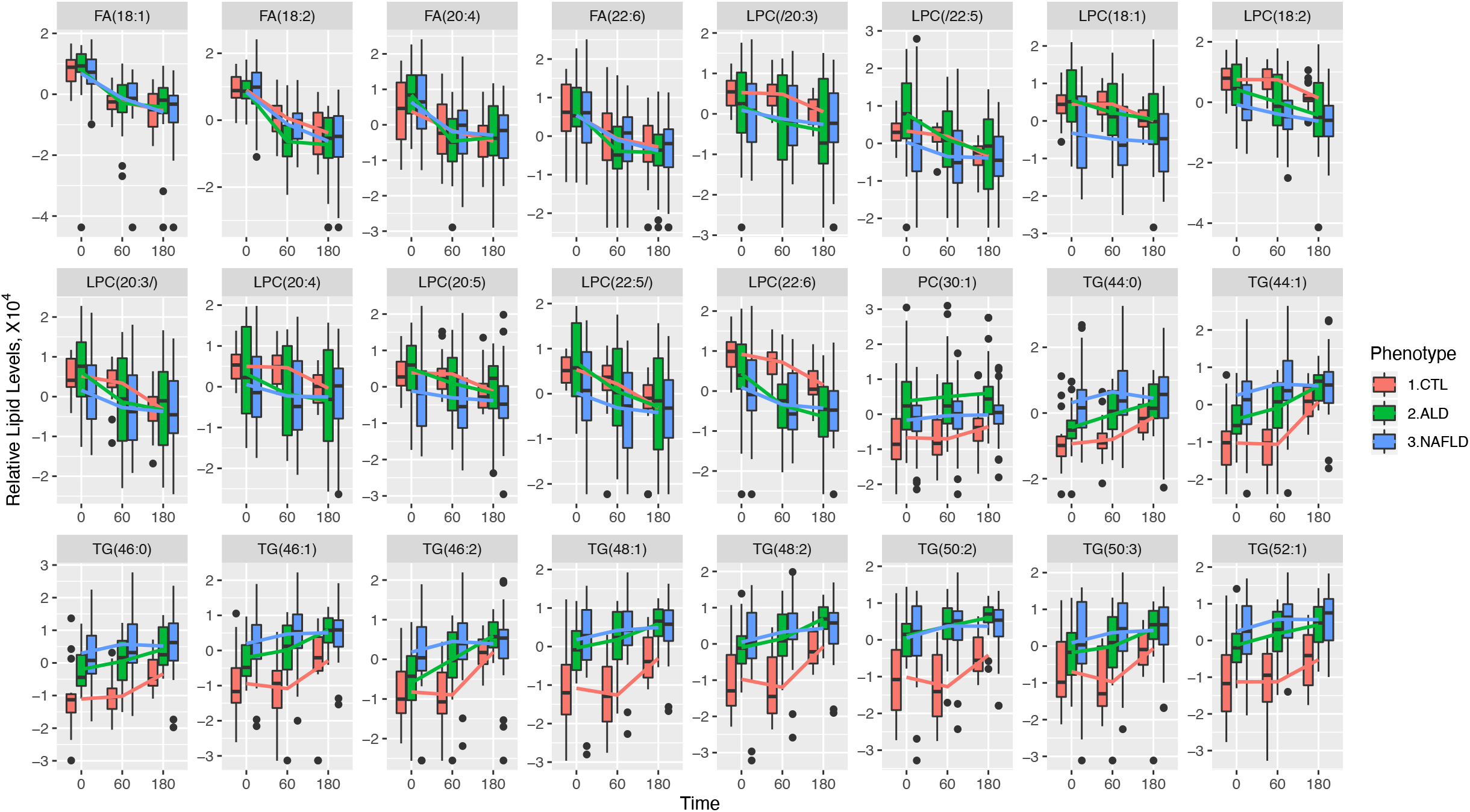
Boxplots showing changes in relative levels of 24 selected hepatic vein lipids from Figure 3a, for three liver phenotypes (controls, ALD, and NAFLD). The lines connect the means of boxes.

We then compared changes (estimate coefficient values) between the hepatic phenotypes. Supplementary Figure 1 shows TG molecules increasing more profoundly in healthy controls, but only TG (44:1) showed a significant difference between healthy controls (+73.08 units per minute) and NAFLD (+4.40 units per minute). We observed similar patterns for FFA and LPC lipids across all three hepatic phenotypes.

Additionally, we carried out longitudinal models in each phenotype for all individual lipids. The models summarized in volcano plots can be seen in Figure 5. In healthy controls, hepatic venous levels of 25 lipids changed after alcohol intervention (p<1.98×10^−4^), these included the 24 lipids found from the earlier model. Two FFA and 8 LPC molecules decreased while 14 TG and one PC molecules increased. In ALD, 19 hepatic venous lipids changed in their level, 4 FFA and 4 LPC decreased while 11 TG increased in their levels. In NALFD, only two FFA hepatic lipids decreased after alcohol intervention.

**Figure 5.**
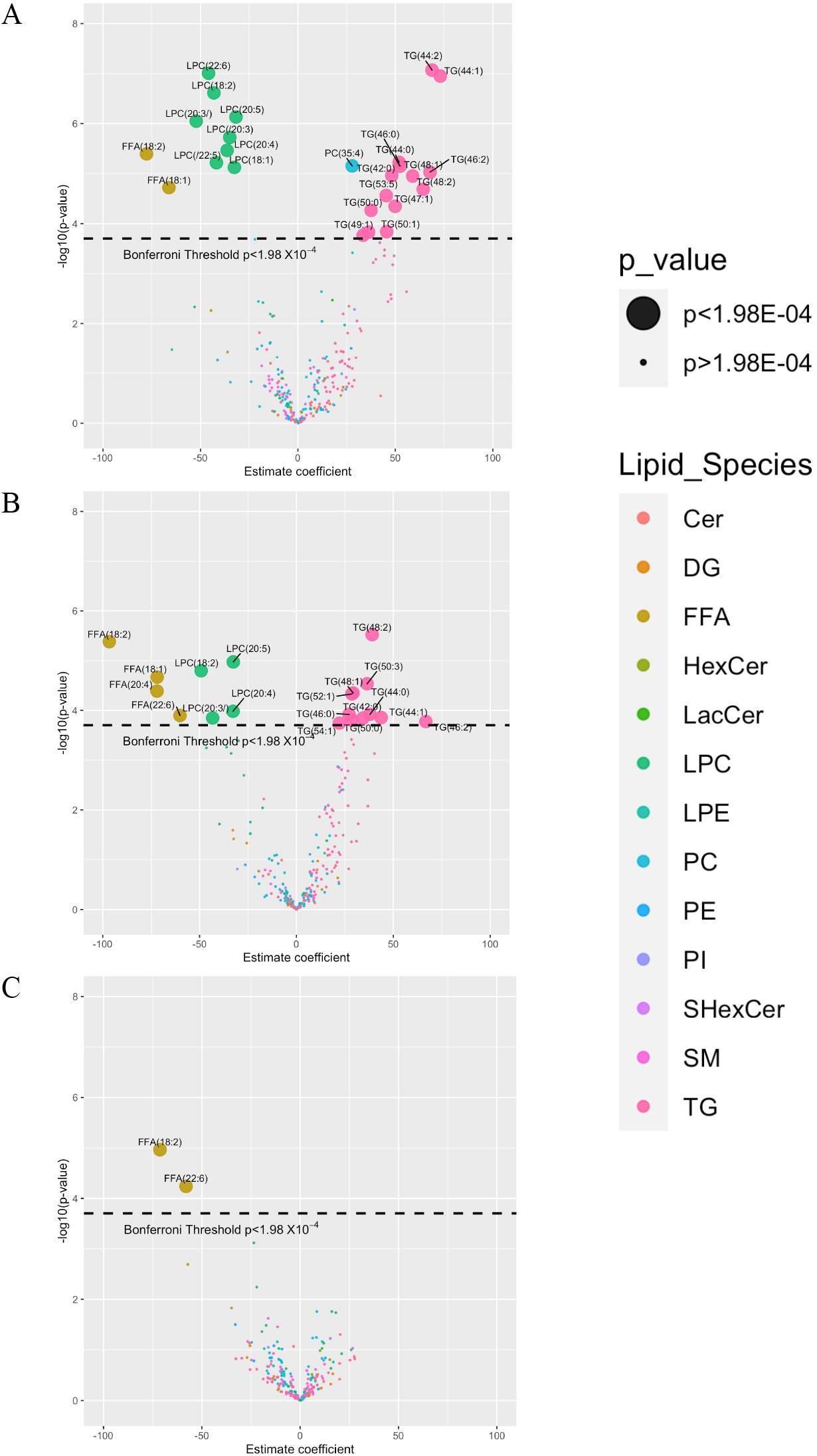
Volcano plot showing changes in levels (estimate coefficients) for 252 lipids from hepatic vein after alcohol intervention in (A) healthy controls, (B) ALD participants and (C) NAFLD participants.

### Single lipid levels in hepatic and systemic venous blood before and after alcohol intervention

Baseline levels of the four FFAs (oleic acid (18:1), linoleic acid (18:2), arachidonic acid (20:4), and docosahexaenoic acid (22:6)) were lower in the hepatic vein than in the systemic vein (Supplementary Table 4 and Figure 6). No single lipid levels changed at a significantly different rate (estimate coefficient) in hepatic and systemic venous blood after alcohol intervention.

**Figure 6.**
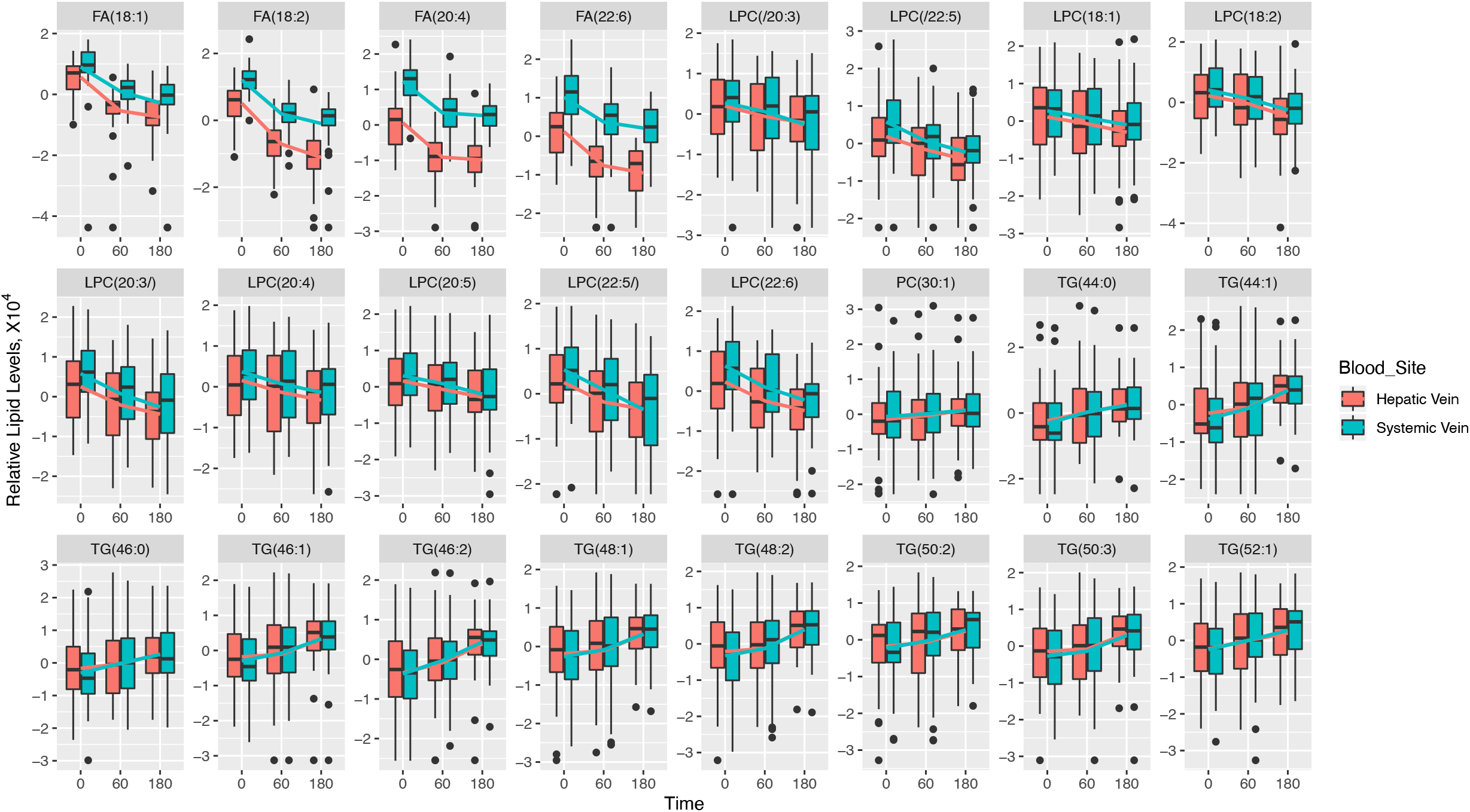
Boxplots showing changes in relative levels of 24 selected lipids from Figure 3a, for hepatic vein vs. systemic vein. The lines connect the means of boxes.

### ALD versus NALFD liver lipidomics

Liver biopsies were collected from ALD and NALFD participants after alcohol intervention and liver lipidomic profiles were compared to identify different lipids levels between the two hepatic phenotypes. Logistic regression model showed no lipid being different between the two hepatic phenotypes at Bonferroni p<1.58×10^−4^ level (Figure 7).

**Figure 7.**
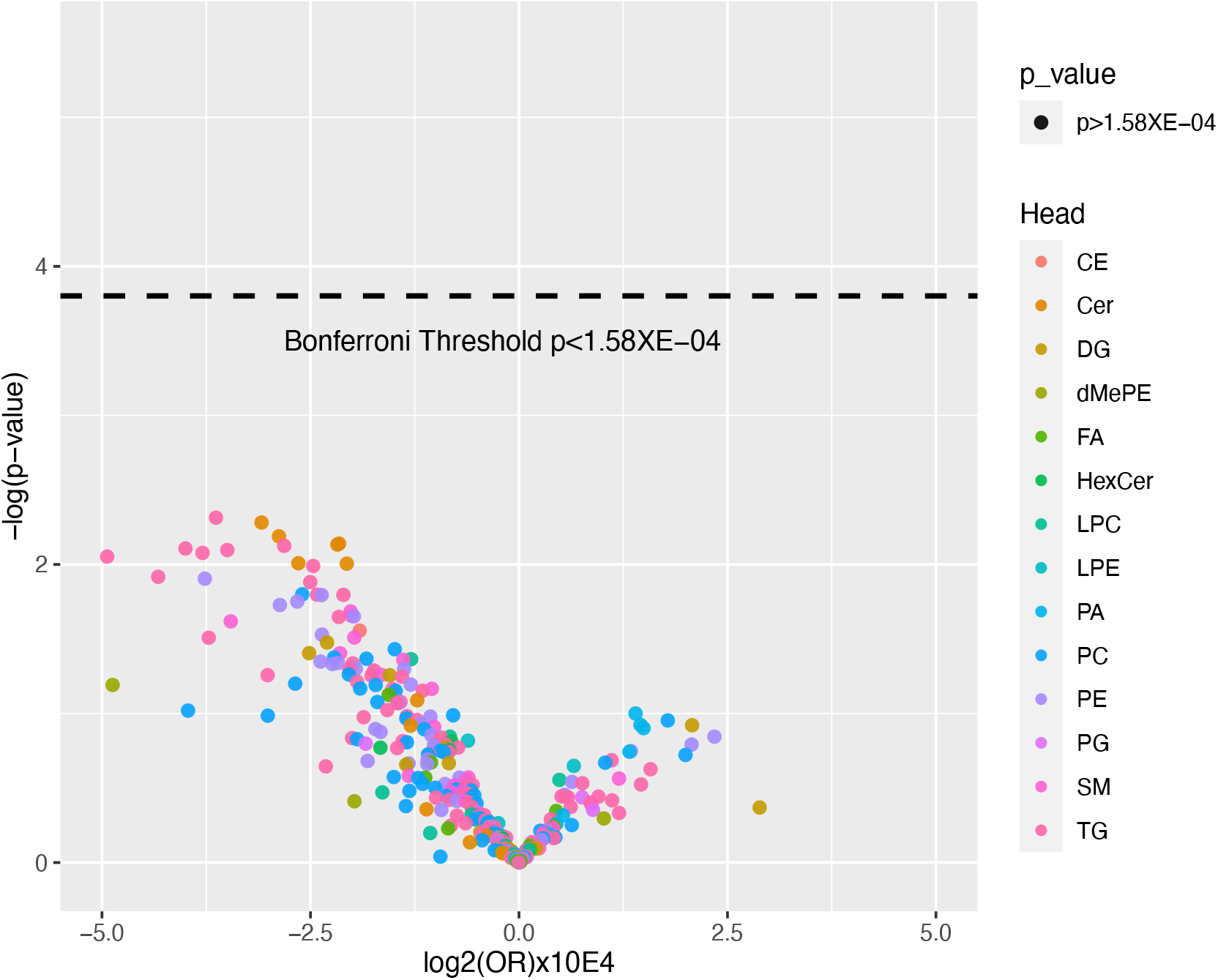
Volcano plot showing differences in levels (OR) for 316 lipids from ALD and NAFLD liver tissue samples from the participants who had undergone alcohol intoxication.

## Discussion

In this first investigation in humans of how acute alcohol intoxication affects the circulating hepatic and systemic lipidome, we found that alcohol caused significantly reduced levels of LPC and FFA, and increased TG levels. Interestingly, FFA changes were driven by two MUFAs and two PUFAs. The FFAs were significantly lower in hepatic blood than in systemic venous blood, probably indicating hepatic uptake. The uptake was similar in ALD, NAFLD, and healthy controls. We observed a corresponding decrease in the levels of LPCs containing the same MUFA and PUFA chains, but only in ALD and healthy controls. Similarly, the increase in TGs was restricted to healthy controls and ALD, indicating a suppressed or altered TG synthesis in NAFLD during acute alcohol intoxication. The TGs that increased were mostly unsaturated or monounsaturated indicating *de novo* lipogenesis.

Our data show a decrease in circulating LPC after alcohol intake. We hypothesize that alcohol increases hepatic LPC uptake which, together with FFA overload, leads to lipoapoptosis and alcoholic steatohepatitis (Figure 8). The role of LPC in the pathogenesis of ALD is not yet known, but intravenous LPC injections induced lobular hepatitis in mice,^12^ and intracellular LPCs were shown to cause caspase activation and generation of oxidative stress resulting in lipoapoptosis of hepatocytes.^12, 13^ Human lipidome profiling of NAFLD liver tissues has revealed that livers with steatohepatitis contain increased amount of LPC compared to livers without steatohepatitis.^14^ Here, we were unable to compare NAFLD liver lipid profiles to those without steatohepatitis because of the absence of liver tissue samples from healthy controls. Lower circulating LPC levels are a marker of poor prognosis in participants with NAFLD,^15, 16^ in agreement with an inverse relationship between circulating and intrahepatic LPC levels. LPC species that decreased after alcohol intervention in our study contained oleic acid (FFA18:1), linoleic acid (FFA18:2), arachidonic acid (FFA20:4), and docosahexaenoic acid (FFA22:6). While the 18-carbon chain are abundant FAs, polyunsaturated FAs are twenty times less abundant in circulation.^17^ Previous studies have demonstrated that in a fasting state, the level of FFAs is lower in hepatic than systemic venous blood indicating hepatic extraction,^18, 19^ and that acute alcohol intoxication induces a rapid drop in the systemic circulating FFA in healthy people and people with ALD and NAFLD-related diseases (type 2 diabetes and hypertriglyceridemia).^20-24^ Our study also observed lower FFA levels, confirming these observations, and adds two further novel findings in humans.

**Figure 8.**
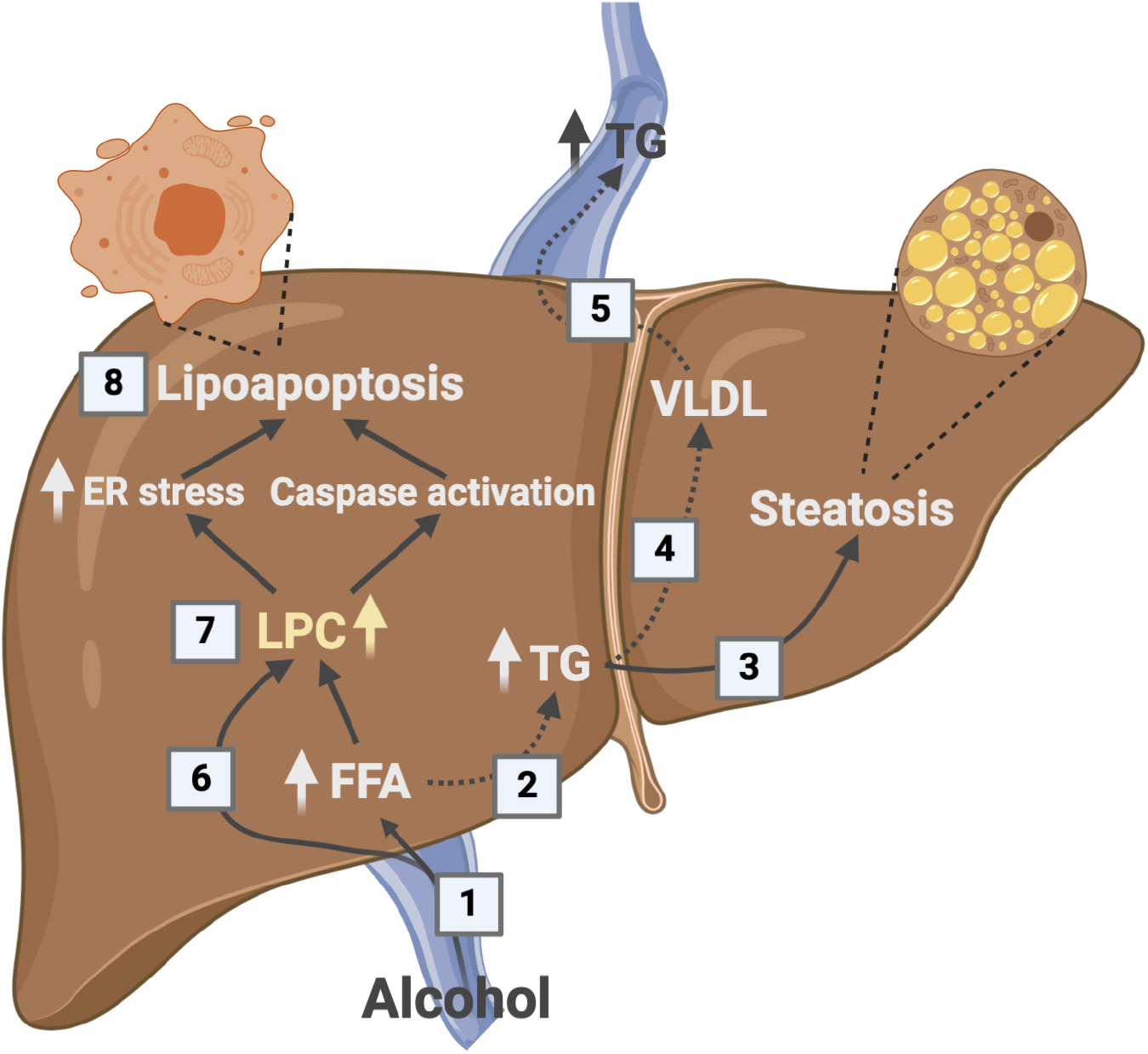
A model of hepatic lipid metabolism after alcohol intake. 1) Alcohol increases hepatic FFA uptake. 2) FFAs can be hydroxylated by CYP2E1 or can be esterified to TGs and either be 3) stored in the hepatocyte leading to steatosis or 4) packaged in VLDL and 5) secreted to the circulation. 6) LPCs can be actively taken up by the liver. Together LPCs and FFAs are the end products of PLA_2_ metabolism. 7) Increased levels of intracellular LPCs may lead to FFAs via lysophospholipase D (LysoPLD) catalysis or to endoplasmic stress and caspase activation that 8) induce lipoapoptosis.^12^ Alcohol intoxication does not lead to increased levels of circulating TGs in NAFLD, which might indicate that at least one of the dashed arrows is suppressed. This may explain the increased TG storage and alcohol susceptibility reported in individuals with metabolic syndrome. FFA, Free fatty acid; ER, endoplasmic reticulum; LPC, lysophosphatidylcholine; TG, triglycerides; VLDL, very-low-density lipoprotein.

Firstly, we observed that the drop in circulating FFAs was mainly driven by mono-(MUFA) and poly-unsaturated (PUFA) FFAs, i.e. oleic acid, linoleic acid, arachidonic acid and docosahexaenoic acid. Our study is in accordance with prior experimental studies where alcohol intake had a profound effect on hepatic fatty acids with 18-carbon acyl chain such as oleic acid and linoleic acid, as well as arachidonic acid and docosahexaenoic acid.^25-28^ These studies were based on mouse, rat, and zebrafish models, and now our findings confirm the same effect in humans. However, the role of unsaturated FFA in fatty liver diseases is unclear and of great importance as the uptake is not observed in the *de novo* synthesized TG composition. Numerous ALD animal studies have documented a damaging effect of dietary unsaturated FFA on alcohol-induced liver injury.^29, 30^ One proposed explanation is that alcohol induces oxidation of unsaturated FFA and that these oxidized unsaturated FFA may contribute to the hepatic steatosis and inflammation seen in ALD.^31^ On the other hand, unsaturated FFA, namely omega-3 PUFAs, have been suggested to play a protective role in NAFLD by reducing hepatic inflammation and TG accumulation, but clinical studies investigating PUFA treatment on NAFLD have led to conflicting results.^32^ Our results show that baseline levels and the acute effect of alcohol on unsaturated FFA are similar in healthy controls, ALD, and NAFLD. Therefore, if unsaturated FFA are involved in development or progression of liver disease, it must be related to either the frequent exposure of alcohol in ALD, or to a change in the subsequent hepatic processing of FFA. Interestingly, the FFA species which changed were same fatty acid chains seen in LPCs, suggesting a profound effect of alcohol on these specific fatty acid chains across two lipid species.

Secondly, we observed that the hepatic uptake of FFAs remained constant despite the alcohol-induced drop in circulating FFAs. This is noteworthy because hepatic FFA β-oxidation is inhibited during alcohol metabolism.^9^ Alcohol-induced suppression of liver FFA β-oxidation in combination with a continued high hepatic uptake of FFAs must lead to an overload of FFAs in the liver. This FFA excess in the liver is metabolized for TG synthesis and released to the circulation. This happened in ALD and healthy controls where circulating TG increased in response to alcohol. In contrast, TG levels remained unaffected after alcohol intake in NAFLD. A suppressed capacity to convert excess hepatic FFA to TG during alcohol intoxication has also been reported in participants with type 2 diabetes and hypercholesterolemia.^21, 24^ A lack of TG synthesis in response to alcohol could indicate that NAFLD either have suppressed hepatic capacity of FFA esterification to TGs, or impaired hepatocyte secretion of TGs. Such impaired ability to remove FFAs from the liver could explain why binge drinking is particularly harmful in individuals with metabolic syndrome.^6^ This mechanism may help explain why high BMI and alcohol have synergistic effect on the risk of liver-related death in the general population.^7^

The pathogenesis of NAFLD is closely related to dyslipidemia.^33^ In line with that, the NAFLD phenotype was significantly different to the ALD and healthy control phenotypes with respect to fasting levels of TG and LPC. However, the levels of TG and LPC in ALD participants changed towards a lipid profile more similar to NAFLD after alcohol intake. Therefore, we speculate that during alcohol intoxication, chronic alcohol misusers with ALD have a functional circulating lipid profile similar to NAFLD. To support this observation of the circulating lipid levels in ALD changing towards those in NALFD, we also looked at liver lipid profiles at 240 minutes in these two hepatic phenotypes which had undergone alcohol intervention and found no difference in lipid profiles between ALD and NAFLD livers after alcohol intoxication.

Limitations of the study warrant consideration. Firstly, it should be emphasized that all investigations in the present study were performed in the fasting state, and lipid dynamics with concurrent food intake may thus be different. Secondly, 38 participants (out of 39) were of European ethnicity. Thirdly, our investigation on blood lipid profiles does not accurately reflect alcohol-induced lipid changes in liver tissue since liver biopsy was acquired only after the intervention. Previous studies have investigated the arterial-hepatic venous difference while here the systemic-hepatic venous difference is compared. ^18, 19^ For ethical and practical reasons the systemic venous blood was collected from the jugular vein catheter to avoid introducing an arterial catheter. There are differences between systemic venous blood and arterial blood, as the blood composition with regards to lipids changes minimally when going through the heart and lungs, and our results are in line with previous study investigating arterial-hepatic venous differences. ^18, 19^ It should be noted, that alcohol inhibits adipose tissue lipolysis and thereby decrease FFA secretion into circulation, which could in partly explain the reduction of FFAs followed by acute alcohol intoxication.^20^ However, we observed that the systemic-hepatic venous FFA difference remained constant during alcohol intoxication, which indicates that hepatic extraction contributes to the decrease of FFAs during alcohol intoxication. Another limitation was the small sample size of liver biopsies, collected in ALD and NAFLD, and only after alcohol intervention hence unable to evaluate changes in liver lipid levels before/after alcohol intervention.

Despite these limitations, there are strengths to this study. We investigated how alcohol intoxication impacts circulating lipid profiles in humans across three distinct hepatic phenotypes. Previous studies have mainly investigated acute alcohol intake in young and healthy individuals. In contrast, our participants were middle-aged with early-stage liver disease and thereby more representative for studying the pathogenesis of chronic fatty liver disease. Furthermore, the comprehensive lipidome analysis enabled us to study lipid classes not previously investigated and indeed reveal that circulating LPCs and specific fatty acid chains are also key in the metabolism of acute alcohol intake.

To conclude, alcohol intoxication induces rapid changes in the profile of circulating lipids. Alcohol has a profound effect on specific LPCs and fatty acids, oleic acid, linoleic acid, arachidonic acid and docosahexaenoic acid. It is likely that these lipids are extracted by the liver during alcohol intoxication. This may play a central role in the pathogenesis of ALD, and the role of LPC in the pathogenesis of ALD should be further explored. This metabolic response to alcohol is suppressed in participants with NAFLD, and that may explain why alcohol is particularly harmful in individuals with metabolic syndrome. Such findings underline the presence of metabolic dysfunction in NAFLD and may fuel the debate of changing the nomenclature to metabolic-associated fatty liver disease (MAFLD).^34, 35^

## Methods

### Participants

Three hepatic phenotypes matched on age were included in this project: I) healthy controls with no sign of liver disease; II) individuals with ALD, III) individuals with NAFLD. Potential participants received written and oral information followed by a pre-investigation consultation with a physician involved in the study. All investigations were performed at Odense Liver Research Centre. The study was approved by the Ethical Committee of Southern Denmark (S-20160083), registered at ClinicalTrials.gov (Identifier: NCT03018990), and complied with the Declaration of Helsinki Declaration of Helsinki – Ethical Principles for Medical Research Involving Human Subjects. All participants signed an informed consent.

Participants were recruited through the Odense Liver Research Centre with inclusion criteria of age 18-75 years, bodyweight >50 kg, capable to be abstinent for 48 hours prior to the investigations and signed informed consent. Individuals with ALD should be regularly drinking and have a prior or ongoing heavy alcohol intake (males >36 g alcohol/day, females >24 g alcohol/day), biopsy-proven liver fibrosis on a previous liver biopsy, and histological features in keeping with ALD. No participants with ALD were abstinent from alcohol or had a desire to become abstinent from alcohol. Individuals with NAFLD should have biopsy-proven liver fibrosis, histological features in keeping with NAFLD, and no history of heavy alcohol intake. Healthy controls should have normal liver stiffness, BMI<30 kg/m^2^, normal biochemistry, and no history of heavy alcohol intake. General exclusion criteria were cirrhosis on a previous liver biopsy, competing liver disease of other etiology, insulin-dependent diabetes mellitus, pregnancy or breastfeeding, antibiotic use within the previous 4 weeks, and all types of cancer.

### Investigations

The investigations were initiated in the morning, after eight hours fasting and minimum 48 hours of abstinence from alcohol. Under local anesthesia, a catheter was placed in a hepatic vein via the right jugular vein and inferior vena cava for sampling hepatic venous blood.^36^ Another catheter was placed in the jugular vein for systemic blood sampling. X-ray was used to ensure correct placement of the catheters prior to blood sampling.

### Alcohol intervention

Ethanol was instilled through a nasogastric tube and was 40% pure ethanol in 9 mg/ml NaCl produced at the hospital pharmacy. Participants received a dose of 2.5 ml of 40% ethanol per kg body weight, instilled over 30 minutes by infusion pump. To avoid severe intoxication in participants with high body fat percentage, the dose was adjusted by 0.5 ml for each kg body weight encountered from BMI above 25 kg/m^2^.

### Blood sampling

Blood was sampled simultaneously from a hepatic vein and the right external jugular vein at baseline (time 0, just before alcohol intervention), after 60 minutes, and after 180 minutes. The Department of Biochemistry and Pharmacology at Odense University Hospital analyzed serum alcohol concentrations and routine biochemistry according to standard operating procedures. Blood samples for lipidome analysis were collected in lithium heparin tubes chilled in crushed ice and water. Within one hour after sampling, we centrifuged and pipetted the samples to secondary tubes and stored them at - 80 °C. A flowchart of the clinical set-up can be found in Figure 1.

### Liver biopsy sampling

Transjugular liver biopsies were performed using Tru-Cut® biopsy needle in participants with ALD and NAFLD 240 minutes after alcohol instillation. First piece of liver tissue from each participant was immediately stored in formalin 4% and send for histopathological assessment and scored according to the NASH Clinical Research Network system with regards to fibrosis, steatosis, inflammation and ballooning.^37, 38^ The second piece of tissue was put into dry tubes and stored immediately at −80 °C until lipidome analysis.

### Lipidome analysis

Sample preparation for lipidomic analysis has been described elsewhere^39, 40^ and is explained in detail in Supplementary Methods 1. Briefly, plasma sample (10 μL) was mixed with 10 μL 0.9% w/v NaCl(aq) and internal standards containing 120 μL of chloroform/methanol (2:1) mixture. The lipid containing chloroform was analyzed using ultra-high-performance liquid chromatograph coupled with quadrupole time-of-flight mass spectrometer (UHPLC-QTOFMS). Samples were analyzed in a randomized order, with quality control (QC) pooled plasma samples at regular intervals throughout the run (n= 30 for both positive and negative ionization).

For liver tissue, the needle biopsy sample was first converted into powered forms by crushing samples using Covaris CryoPrep impactor CP02. Approximately 2 mg of powered tissues sample was used for lipid extraction (exact mass was measured and used for post-processing normalization step later). The sample was mixed with internal standards containing 480 μL of chloroform/methanol (2:1) mixture. The lipid containing chloroform was analyzed in the same manner as blood samples.

The lipidomics data were pre-processed with MZmine2^41^, and then lipid features were normalized to internal standards and log transformed. Liver samples were further normalized to the exact weight used during lipid extraction. We excluded lipid features with >20% missingness across all samples and relative standard deviation (RSD) values >20% across QC samples. The lipid features were then standardized (scaled) to have a mean of 0 and a standard deviation of 1×10^4^. The data were cross-matched with an in-house library where 252 lipids from 13 different lipid classes were identified in blood samples while 316 lipids from 14 different lipid classes were identified in liver tissue samples at level 1and 2 lipid identification levels.

### Statistical analysis

Participants’ baseline characteristics are reported as mean and standard deviations (SD), counts and proportion, or medians and interquartile range (IQR) depending on the distribution. For blood lipidomic analysis, we applied longitudinal mixed effect regression models with participants as random effect to investigate changes in circulating lipid contents after alcohol intake.

First, we explored the average level of 13 distinct lipid classes from hepatic venous blood to identify which lipid classes changed in level over time after alcohol intervention. For this, we derived average levels from each lipid class and built mixed models between levels of 13 lipid classes and three different fixed effects: (1) ‘time’, (2) ‘time*phenotype’, and (3) ‘time*blood site’. Fixed effect of ‘time’ allowed us to investigate changes in circulating lipid content after alcohol intervention. From these models we derived estimate coefficient values (representing changes in relative levels per minute) and their corresponding p-values. Fixed effect of ‘time*phenotype’ allowed us to see whether levels of 13 lipid classes were different between phenotypes at baseline (before alcohol intervention) and whether estimate coefficient values of lipid classes differed between the three phenotypes. Fixed effect of ‘time*blood site’ allowed us to see whether baseline levels and estimate coefficient values of the 13 lipids classes differed between the two blood vein sites (systemic vs. hepatic).

From the 13 lipid classes, we investigated 252 individual lipid species. We used the same approach as above where fixed effects were (1) ‘time’, (2) ‘time*phenotype’, and (3) ‘time*blood site’. In all models, random effect was the individual participant. We report the significant lipids after Bonferroni correction for multiple testing. Mixed models were carried out in R 3.6.0 using the ‘nlme’ package.^42^

Lipid levels (n=316, 14 lipid classes) from liver biopsy samples were available from 14 ALD and 15 NAFLD participants after alcohol intervention. We investigated effect of alcohol intervention between ALD and NAFLD liver lipids by utilizing logistic regression model where age of participants at the time of sampling was adjusted. Regression model was carried out in R 3.6.0. A schematic workflow of the of the lipidome data analysis is shown in Figure 1.

## Data Availability

Anonymised data on request.

## Abbreviations used in this paper

ALD: alcohol-related liver disease
ALT: alanine aminotransferase
ANOVA: analysis of variance
AST: asparagine aminotransferase
BMI: body mass index
Cer: ceramide
CTL: healthy control
DG: diglyceride
FFA: free fatty acid
GGT: gamma-glutamyl transferase
HbA1c: hemoglobin A1c
HDL: high-density lipoprotein
HexCer: hexosylceramide
HOMA-IR: Homeostatic Model Assessment of Insulin Resistance
IQR: interquartile range
LacCer: lactosylceramides
LCAT: lecithin:cholesterol acyltransferase
LPC: lysophosphatidylcholine
LPE: lysophosphatidylethanolamine
INR: international normalized ratio
LDL: low-density lipoprotein
NAFLD: non-alcoholic fatty liver disease
PC: phosphatidylcholine
PE: phosphatidylethanolamine
P-glucose: plasma glucose
PI: phosphatidylinositol
PLA2: phospholipase A2
QC: quality control
SD: standard deviation
SHexCer: sulfatides hexosylceramide
SM: sphingomyelin
TE: transient elastography
TG: triglyceride

## Acknowledgements

The authors thank Mie Balle Hugger, Annette Fialla, Annette Nielsen, Lotte Kappen and Jane Brudvig-Lauridsen and the entire staff at Odense Liver Research Centre; Mette Andreasen, Anette Tyrsted and Lea Grip from Open Patient data Explorative Network; Professor Peter Rossing at Steno Diabetes Centre Copenhagen; Associate professor Claire Gudex from University of Southern Denmark; and the MicrobLiver consortium partners.

## Author contributions

MI, BSM, TH and AK conceptualized the study; MI, MK, BSM, CDH, NT and KT collected data for the study; MI, MK, TS, KT and CLQ performed the data analyses; MI, MK, MT, CLQ and AK drafted the manuscript; all authors contributed to the manuscript with important intellectual content and approved the final version.

### Grant support

Challenge Grant “MicrobLiver”, NNF15OC0016692, Novo Nordisk Foundation.

