## Supplementary material for "Comprehensive lipidomics reveals reduced hepatic lipid turnover in NAFLD during alcohol intoxication"

#### Alcohol intoxication and hepatic lipid metabolism: Lipidome analysis of hepatic and systemic vein blood in humans

### Supplementary Methods 1. Lipidomics

#### Sample preparation

Sample preparation has been described elsewhere.(Tofte 2019 & Bowden 2017) 10µL of plasma sample was added to a 2mL Eppendorf tube. To the samples were added 10µL 0.9% w/v NaCl(aq), 28µL chloroform/methanol (2:1) containing 14 internal standards (10µg/mL for all), and 92µL chloroform/methanol (2:1). The internal standards used were 2-diheptadecanoyl-sn-glycero-3-phosphoethanolamine (PE; 17:0/17:0), N-heptadecanoyl-D-erythro-sphingosylphosphorylcholine (SM; d18:1/17:0), N-heptadecanoyl-D-erythro-sphingosine (Cer; d18:1/17:0), 1,2-diheptadecanoyl-sn-glycero-3-phosphocholine (PC; 17:0/17:0), 1-heptadecanoyl-2-hydroxy-sn-glycero-3-phosphocholine (LPC; 17:0), 1,2-Dimyristoyl-sn-glycero-3-phospho(choline-d13) (PC; 14:0/d13), Tripalmitin-1,1,1-13C3 (TG; (16:0/16:0/16:0)-13C3), Trioctanoin-1,1,1-13C3 (TG; (8:0/8:0/8:0)-13C3), 1-palmitoyl-d31-2-oleoyl-sn-glycero-3-phosphocholine (PC; 16:0/d31/18:1), 1,2-diheptadecanoyl-sn-glycero-3-phospho-(1'-rac-glycerol) (PG; 17:0/17:0), 1,2-diheptadecanoyl-sn-glycero-3-phospho-L-serine (PS; 17:0/17:0), 1,2-diheptadecanoyl-sn-glycero-3-phosphophate (PA; 17:0/17:0), Trinonadecanoin (TG; 19:0/19:0/19:0), and Tripentadecanoin (TG; 15:0/15:0/15:0). The mixture was vortexed and centrifuged (1000g, 4 minutes, 4°C). The lipid containing chloroform phase (lower level, 30µL) was extracted to a glass HPLC vial containing a 400µL glass insert. The samples were stored at −80 °C until analysis on the UHPLC-QTOF-MS system.

#### UHPLC-MS analysis

The UHPLC system used for lipidomics was a 1290 Infinity system (Agilent, Santa Clara, California, USA). Separations were performed on an ACQUITY UPLC® BEH C18 column (2.1 mm × 100 mm,

1.7 µm) along with a precolumn (BEH Shield RP18, 2.1 × 5 mm, 1.7 µm), both by Waters (Milford, USA). Mobile phase used were $H_2O$ + 1% $NH_4Ac$ (1M) + 0.1% v/v HCOOH (A) and ACN:IPA (1:1, v/v) + 1% NH4Ac + 0.1% HCOOH (B). The gradient was: 0 - 2 min. 35-80% B; 2 - 7 min. 80–100% B; and 7 - 14 min 100% B. This was followed by a 4 min re-equilibration period under the initial conditions (35% B). The flow rate (0.4 mL $min^{-1}$) and the column temperature were maintained at 40°C throughout the run. The injection volume was 1µL.

UHPLC was coupled to a 6550 QTOF-MS interfaced with a dual jet stream electrospray (dual ESI) ion source (Agilent, Santa Clara, California, USA). The drying gas flow was 14 L min−1 (at 193 °C for positive mode and at 185 °C for negative mode). The sheath gas flow was 11 L min−1 (at 379 °C for positive mode and at 345 °C for negative mode). The capillary voltage was 3643V for positive mode and 4300V for negative mode. The nozzle voltage was 1500V for positive mode and 1400V for negative mode. All analyses were acquired using the lock spray setting where Agilent reference mass solution was used as lock mass (positive m/z 121.0509 and 922.0098; negative m/z 119.0363 and 966.0007). Data were collected in centroid mode over the mass range m/z 100 – 1000 with an acquisition time of 0.2 seconds per scan.

Samples were analyzed in a randomized order, with quality control pooled plasma sampled (QC) at regular intervals throughout the run (n= 30 for both positive and negative ionization).

**Data processing**

The acquired lipidomics data was pre-processed with MZmine2 (Pluskal 2010) which performed filtration, peak identification, matching of peaks across samples, and retention time correction.

Positive and negative ionization mode data were extracted separately, then lipid features were normalized to internal standards and log transformed. Lipid features with >20% missingness across all samples and QC relative standard deviation (RSD) values >20% were excluded. Lipid features were then scaled to have a mean of 0 and a standard deviation of $1\times10^4$. The data were cross-matched with an in-house library where 252 lipids from 13 different lipid classes were identified at level 1 and 2. These 13 lipid classes comprised ceramide (Cer), diglyceride (DG), free fatty acid (FFA), hexosylceramide (HexCer), lactosylceramides (LacCer), lysophosphatidylcholine (LPC), lysophosphatidylethanolamine (LPE), phosphatidylcholine (PC), phosphatidylethanolamine (PE), phosphatidylinositol (PI), sulfatides hexosylceramide (SHexCer), sphingomyelin (SM), and triglyceride (TG).

| Fixed Effect Model | Variable | Vein Site | Cer | DG | FFA | HexCer | LacCer | LPC | LPE | PC | PE | PI | SHexCer | SM | TG |
|---|---|---|---|---|---|---|---|---|---|---|---|---|---|---|---|
| 1 | **Time Est.Coef. (95% CI)** | **Hep.** | 1.05 (-4.72~6.83) | -3.42 (-12.75~5.91) | **-39.42 (-50.95~-27.89)** | -0.21 (-6.23~5.81) | 0.38 (-6.92~7.68) | **-14.08 (-21.28~-6.87)** | -4.03 (-15.16~7.09) | -3.51 (-8.83~1.80) | -7.29 (-14.25~-0.34) | -1.90 (-9.19~5.38) | -1.92 (-17.88~14.03) | -4.51 (-11.02~2.00) | 12.00 (2.16~21.84) |
| | **Time Slope p-value** | **Hep.** | $7.17\times10^{-01}$ | $4.68\times10^{-01}$ | **$*1.91\times10^{-09}$** | $9.45\times10^{-01}$ | $9.18\times10^{-01}$ | **$*2.10\times10^{-04}$** | $4.73\times10^{-01}$ | $1.92\times10^{-01}$ | $4.01\times10^{-02}$ | $6.04\times10^{-01}$ | $8.11\times10^{-01}$ | $1.72\times10^{-01}$ | $1.76\times10^{-02}$ |
| 2 | **CTL Est.Coef. (95% CI)** | **Hep.** | 2.02 (-6.66~10.70) | 2.34 (-17.58~22.26) | **-31.02 (-50.13~-11.90)** | -5.55 (-18.83~7.72) | 6.92 (-0.12~13.95) | **-23.96 (-33.88~-14.04)** | -16.17 (-36.38~4.04) | -5.33 (-12.27~1.62) | -5.85 (-19.37~7.67) | 6.75 (-0.63~14.12) | -5.97 (-54.35~42.42) | -8.88 (-21.77~4.02) | **25.20 (11.19~39.22)** |
| | **CTL Slope p-value** | **Hep.** | $6.32\times10^{-01}$ | $8.80\times10^{-01}$ | **$*3.04\times10^{-03}$** | $3.92\times10^{-01}$ | $5.37\times10^{-02}$ | **$*7.02\times10^{-05}$** | $1.01\times10^{-01}$ | $1.25\times10^{-01}$ | $3.76\times10^{-02}$ | $7.06\times10^{-02}$ | $7.99\times10^{-01}$ | $1.66\times10^{-01}$ | **$*1.31\times10^{-03}$** |
| 3 | **ALD (95% CI)** | **Hep.** | 0.06 (-11.65~11.77) | -9.89 (-23.97~4.19) | **-44.36 (-65.28~-23.44)** | -4.80 (-14.64~5.03) | 0.61 (-12.87~14.09) | -20.97 (-34.68~-7.26) | -2.93 (-22.50~16.63) | -4.34 (-14.92~6.23) | -6.10 (-20.05~7.85) | -2.88 (-16.28~10.52) | 0.89 (-18.57~20.36) | -3.52 (-18.17~11.14) | **17.52 (6.33~28.70)** |
| | **ALD Slope p-value** | **Hep.** | $9.91\times10^{-01}$ | $1.61\times10^{-01}$ | **$*1.74\times10^{-04}$** | $3.25\times10^{-01}$ | $9.27\times10^{-01}$ | $4.08\times10^{-03}$ | $7.61\times10^{-01}$ | $4.07\times10^{-01}$ | $3.77\times10^{-02}$ | $6.63\times10^{-01}$ | $9.26\times10^{-01}$ | $6.27\times10^{-01}$ | **$*3.38\times10^{-03}$** |
| 4 | **NAFLD Est.Coef. (95% CI)** | **Hep.** | 1.34 (-8.29~10.96) | -1.22 (-18.17~15.74) | **-40.41 (-61.08~-19.74)** | 7.64 (-1.97~17.26) | -4.19 (-18.47~10.08) | -1.05 (-12.64~10.53) | 3.03 (-16.44~22.51) | -1.53 (-10.91~7.86) | -9.36 (-19.71~0.99) | -6.76 (-20.82~7.30) | -1.86 (-24.33~20.62) | -2.52 (-9.68~4.64) | -1.95 (-23.56~19.65) |
| | **NAFLD Slope p-value** | **Hep.** | $7.78\times10^{-01}$ | $8.84\times10^{-01}$ | **$*4.02\times10^{-04}$** | $1.15\times10^{-01}$ | $5.53\times10^{-01}$ | $8.54\times10^{-01}$ | $7.52\times10^{-01}$ | $7.41\times10^{-01}$ | $7.45\times10^{-02}$ | $3.33\times10^{-01}$ | $8.67\times10^{-01}$ | $4.77\times10^{-01}$ | $8.55\times10^{-01}$ |
| 5 | **CTLvs. ALD Slope p-value** | **Hep.** | $7.99\times10^{-01}$ | $2.90\times10^{-01}$ | $3.57\times10^{-01}$ | $9.24\times10^{-01}$ | $4.51\times10^{-01}$ | $7.38\times10^{-01}$ | $3.47\times10^{-01}$ | $8.84\times10^{-01}$ | $9.79\times10^{-01}$ | $2.52\times10^{-01}$ | $7.61\times10^{-01}$ | $5.92\times10^{-01}$ | **$3.75\times10^{-01}$** |
| | **CTLvs. ALD Baseline p-value** | **Hep.** | $6.34\times10^{-01}$ | $3.52\times10^{-01}$ | $4.54\times10^{-01}$ | $1.27\times10^{-01}$ | $1.31\times10^{-01}$ | $4.03\times10^{-02}$ | $2.67\times10^{-01}$ | $1.18\times10^{-01}$ | $5.96\times10^{-01}$ | $1.93\times10^{-01}$ | $9.90\times10^{-01}$ | $1.68\times10^{-02}$ | **$*6.43\times10^{-06}$** |
| 6 | **CTL vs. NAFLD Slope p-value** | **Hep.** | $9.19\times10^{-01}$ | $7.82\times10^{-01}$ | $5.19\times10^{-01}$ | $9.48\times10^{-02}$ | $2.23\times10^{-01}$ | $5.94\times10^{-03}$ | $1.79\times10^{-01}$ | $5.46\times10^{-01}$ | $6.68\times10^{-01}$ | $1.36\times10^{-01}$ | $8.60\times10^{-01}$ | $3.40\times10^{-01}$ | **$5.96\times10^{-02}$** |
| | **CTL vs. NAFLD Baseline p-value** | **Hep.** | $6.68\times10^{-01}$ | $4.27\times10^{-01}$ | $5.12\times10^{-01}$ | $1.96\times10^{-01}$ | $1.90\times10^{-01}$ | $2.81\times10^{-02}$ | $2.60\times10^{-01}$ | $2.10\times10^{-01}$ | $4.51\times10^{-01}$ | $3.03\times10^{-01}$ | $9.91\times10^{-01}$ | $2.16\times10^{-02}$ | **$*8.38\times10^{-04}$** |
| 7 | **ALD vs. NAFLD Slope p-value** | **Hep.** | $8.63\times10^{-01}$ | $4.27\times10^{-01}$ | $7.84\times10^{-01}$ | $6.92\times10^{-02}$ | $6.19\times10^{-01}$ | $2.62\times10^{-02}$ | $6.60\times10^{-01}$ | $6.84\times10^{-01}$ | $6.99\times10^{-01}$ | $6.85\times10^{-01}$ | $8.51\times10^{-01}$ | $8.99\times10^{-01}$ | $1.14\times10^{-01}$ |
| | **ALD vs. NAFLD Baseline p-value** | **Hep.** | $6.33\times10^{-01}$ | $8.89\times10^{-01}$ | $6.35\times10^{-01}$ | $3.21\times10^{-01}$ | $2.01\times10^{-01}$ | $6.35\times10^{-01}$ | $5.70\times10^{-01}$ | $8.14\times10^{-01}$ | $7.39\times10^{-01}$ | $4.16\times10^{-01}$ | $4.03\times10^{-02}$ | $9.24\times10^{-03}$ | $2.01\times10^{-01}$ |
| 8 | **Time Est.Coef. (95% CI)** | **Peri.** | -4.95 (-12.66~2.76) | -10.65 (-22.30~1.01) | **-37.91 (-50.00~-25.82)** | -5.63 (-17.74~6.48) | 2.01 (-5.93~9.95) | **-14.84 (-21.11~-8.56)** | 3.16 (-5.12~11.45) | -3.12 (-7.31~1.06) | -1.01 (-6.60~4.58) | -1.53 (-9.33~6.27) | -9.29 (-39.41~20.83) | -4.05 (-11.31~3.20) | **16.53 (8.92~24.14)** |
| | **Time Slope p-value** | **Peri.** | $2.05\times10^{-01}$ | $7.28\times10^{-2}$ | **$*2.15\times10^{-08}$** | $3.58\times10^{-01}$ | $6.16\times10^{-01}$ | **$*1.08\times10^{-05}$** | $4.50\times10^{-01}$ | $1.42\times10^{-01}$ | $7.20\times10^{-01}$ | $6.98\times10^{-01}$ | $5.41\times10^{-01}$ | $2.70\times10^{-01}$ | **$*4.55\times10^{-05}$** |
| 9 | **Peri. vs. Hep. Slope p-value** | **Peri. vs. Hep.** | $2.42\times10^{-01}$ | $3.37\times10^{-01}$ | **$8.56\times10^{-01}$** | $4.28\times10^{-01}$ | $7.69\times10^{-01}$ | $8.67\times10^{-01}$ | $3.05\times10^{-01}$ | $9.08\times10^{-01}$ | $1.66\times10^{-01}$ | $9.47\times10^{-01}$ | $6.60\times10^{-01}$ | $5.05\times10^{-01}$ | $2.56\times10^{-01}$ |
| | **Peri. vs. Hep. Baseline p-value** | **Peri. vs. Hep.** | $6.89\times10^{-01}$ | $4.05\times10^{-01}$ | **$*1.44\times10^{-15}$** | $8.88\times10^{-01}$ | $8.13\times10^{-01}$ | $1.04\times10^{-01}$ | $9.43\times10^{-01}$ | $3.34\times10^{-01}$ | $5.25\times10^{-01}$ | $7.08\times10^{-01}$ | $8.51\times10^{-01}$ | $9.28\times10^{-01}$ | $4.59\times10^{-01}$ |

**Supplementary Table 1. Summary of mixed models investigation association between average levels of lipid classes and 9 different fixed effects.** The fixed effects shown in this table includes (1) Time from hepatic vein, (2) Time in CTL from hepatic, (3) Time in ALD from hepatic, (4) Time in NAFLD from hepatic, (5) Time*(CTL vs. ALD) from hepatic, (6) Time*(CTL vs. NAFLD) from hepatic, (7) Time*(ALD vs. NAFLD) from hepatic, (8) Time from systemic, and (9) Time*(Systemic vs. Hepatic). When fixed effect was a single variable (e.g., Time), estimate coefficient values (change in relative level of lipid specie every minute after alcohol intervention) and their p-values were calculated. When fixed effect was an interaction between variables (e.g., Time*(CTL vs. ALD)), slope p-values representing significant differences between changes in relative levels of lipid species between groups, and baseline p-values representing significant differences between relative levels of lipid species at baseline (time = 0min) were derived.

**Abbreviation**. CTL, Control healthy; ALD, Alcohol related liver disease; NAFLD, Non-alcohol related fatty liver disease; Est. Coef., Estimate coefficient; CI, Confidence interval;, Hep., Hepatic; Peri., Systemic

*Passed Bonferroni procedure $p<3.85\times10^{-03}$.

**Relative changes in lipid levels after alcohol intake**

| Lipid Class | Hepatic vein Estimate Coefficient (95% CI) | | | | Systemic vein Estimate Coefficient (95% CI) | | | |
|---|---|---|---|---|---|---|---|---|
| | **All** | **CTL** | **ALD** | **NAFLD** | **All** | **CTL** | **ALD** | **NAFLD** |
| **Cer** | 1.05 (-4.72~6.83) | 2.02 (-6.66~10.70) | 0.06 (-11.65~11.77) | 1.34 (-8.29~10.96) | -4.95 (-12.66~2.76) | -6.84 (-31.01~17.33) | -5.97 (-17.14~5.20) | -2.74 (-11.70~6.23) |
| **DG** | -3.42(-12.75~5.91) | 2.34 (-17.58~22.26) | -9.89 (-23.97~4.19) | -1.22 (-18.17~15.74) | -10.65(-22.30~1.01) | -11.71 (-38.47~15.05) | -13.30 (-31.44~4.85) | -7.46 (-26.26~11.34) |
| **FFA** | ***-39.42 (-50.95~-27.89)** | ***-31.02 (-50.13~-11.90)** | ***-44.36 (-65.28~-23.44)** | ***-40.41 (-61.08~-19.74)** | ***-37.91 (-50.00~-25.82)** | ***-45.97 (-76.04~-15.89)** | ***-30.67 (-48.95~-12.40)** | ***-39.29 (-58.91~-19.67)** |
| **HexCer** | -0.21 (-6.23~5.81) | -5.55 (-18.83~7.72) | -4.80 (-14.64~5.03) | 7.64 (-1.97~17.26) | -5.63 (-17.74~6.48) | -14.13 (-55.78~27.53) | -2.21 (-15.92~11.50) | -3.15 (-16.30~10.00) |
| **LacCer** | 0.38 (-6.92~7.68) | 6.92 (-0.12~13.95) | 0.61 (-12.87~14.09) | -4.19 (-18.47~10.08) | 2.01 (-5.93~9.95) | 5.85 (-14.77~26.46) | -2.83 (-16.97~11.32) | 3.96 (-6.26~14.18) |
| **LPC** | ***-14.08 (-21.28~-6.87)** | ***-23.96 (-33.88~-14.04)** | -20.97 (-34.68~-7.26) | -1.05 (-12.64~10.53) | ***-14.84 (-21.11~-8.56)** | ***-15.50 (-30.02~-0.98)** | -20.52 (-31.86~-9.18) | -9.09 (-17.96~-0.23) |
| **LPE** | -4.03 (-15.16~7.09) | -16.17 (-36.38~4.04) | -2.93 (-22.50~16.63) | 3.03 (-16.44~22.51) | 3.16 (-5.12~11.45) | -2.63 (-15.62~10.35) | -2.72 (-18.32~12.89) | 12.52 (-1.59~26.62) |
| **PC** | -3.51 (-8.83~1.80) | -5.33 (-12.27~1.62) | -4.34 (-14.92~6.23) | -1.53 (-10.91~7.86) | -3.12 (-7.31~1.06) | -2.82 (-13.46~7.83) | -5.47 (-11.50~0.55) | -1.13 (-8.03~5.77) |
| **PE** | -7.29 (-14.25~-0.34) | -5.85 (-19.37~7.67) | -6.10 (-20.05~7.85) | -9.36 (-19.71~0.99) | -1.01 (-6.60~4.58) | -2.35 (-14.21~9.51) | 0.41 (-9.43~10.24) | -1.43 (-10.64~7.78) |
| **PI** | -1.90 (-9.19~5.38) | 6.75 (-0.63~14.12) | -2.88 (-16.28~10.52) | -6.76 (-20.82~7.30) | -1.53 (-9.33~6.27) | -9.47 (-34.33~15.39) | -1.55 (-11.24~8.14) | 3.79 (-5.92~13.50) |
| **SHexCer** | -1.92 (-17.88~14.03) | -5.97 (-54.35~42.42) | 0.89 (-18.57~20.36) | -1.86 (-24.33~20.62) | -9.29 (-39.41~20.83) | -60.81 (-166.45~44.82) | 1.85 (-26.22~29.93) | 14.66 (-14.49~43.82) |
| **SM** | -4.51 (-11.02~2.00) | -8.88 (-21.77~4.02) | -3.52 (-18.17~11.14) | -2.52 (-9.68~4.64) | -4.05 (-11.31~3.20) | 3.10 (-15.06~21.26) | -7.57 (-21.27~6.14) | -5.54 (-14.28~3.19) |
| **TG** | 12.00 (2.16~21.84) | ***25.20 (11.19~39.22)** | 17.52 (6.33~28.70) | -1.95 (-23.56~19.65) | ***16.53 (8.92~24.14)** | ***12.88 (-7.02~32.77)** | 19.49 (6.49~32.50) | 16.20 (5.77~26.63) |

**Supplementary Table 2. Summary of mixed models showing relative changes from baseline in lipid class levels after alcohol intake.** In this table, estimate coefficient values (change in relative level of lipid every minute after alcohol intervention) and their p-values are shown.
Abbreviation. CI, Confidence interval; CTL, Control healthy; ALD, Alcohol-related liver disease; NAFLD, Non-alcoholic steatohepatitis
*Significant association at Bonferroni procedure $p<3.85\times10^{-03}$ level.

| Blood sampling (t = 0 minute) | | | | | | | | |
|---|---|---|---|---|---|---|---|---|
| **Lipid Class** | **Average lipid level in Hepatic vein (S.D.), ×10$^{02}$** | | | | **Average lipid level in Systemic vein (S.D. ), ×10$^{02}$** | | | |
| | **All** | **CTL** | **ALD** | **NAFLD** | **All** | **CTL** | **ALD** | **NAFLD** |
| **Cer** | -1.19(59.42) | 4.65(65.43) | -1.47(48.84) | -4.82(67.67) | 2.85(60.77) | 2.80(78.53) | 6.25(53.21) | -0.28(58.38) |
| **DG** | 2.99(47.00) | -16.20(47.74) | 7.66(34.32) | 11.43(55.47) | 5.56(63.44) | -9.15(56.23) | 14.23(64.86) | 7.27(68.95) |
| **FFA** | *12.92(45.06) | 5.64(48.46) | 21.41(28.90) | 9.85(55.72) | 80.21(59.73) | 104.92(61.19) | 62.15(48.17) | 80.60(66.04) |
| **HexCer** | 2.67(73.43) | 28.56(84.26) | 19.87(43.33) | -30.64(79.78) | 2.04(74.75) | 12.05(107.17) | 20.77(48.21) | -22.11(67.66) |
| **LacCer** | -6.13(77.93) | 36.26(86.41) | -23.60(61.19) | -18.07(80.40) | 3.08(87.52) | 36.93(79.24) | -6.79(79.96) | -10.28(98.54) |
| **LPC** | 5.13(84.72) | 46.14(28.05) | 18.49(79.81) | -34.68(99.92) | 22.85(75.97) | 46.26(34.65) | 36.46(80.27) | -5.46(86.45) |
| **LPE** | 3.59(79.80) | 23.27(63.34) | 21.38(82.89) | -26.13(82.39) | -4.10(72.56) | 9.76(43.95) | 19.72(74.01) | -35.57(78.72) |
| **PC** | -1.36(50.08) | 16.01(42.13) | 1.72(34.92) | -15.81(63.92) | 5.67(45.90) | 22.69(42.00) | 7.67(36.15) | -7.55(54.53) |
| **PE** | 0.49(57.65) | 10.95(50.52) | -4.35(73.09) | -1.98(48.08) | 4.65(49.58) | 8.60(34.05) | 8.21(69.14) | -1.32(37.77) |
| **PI** | -1.10(64.17) | -21.25(53.42) | 18.84(52.61) | -6.26(77.83) | 1.05(70.80) | -9.17(58.06) | 22.64(50.93) | -12.29(91.11) |
| **SHexCer** | 7.26(74.85) | -11.02(113.39) | 25.66(50.75) | 2.28(62.83) | -9.96(90.26) | -14.82(119.57) | 12.23(66.70) | -27.43(89.20) |
| **SM** | -0.38(75.77) | 54.65(85.80) | -41.50(54.97) | 1.32(64.72) | 7.44(70.69) | 34.05(74.81) | -20.94(69.22) | 16.18(64.21) |
| **TG** | -13.44(76.09) | -67.27(43.10) | -20.68(47.79) | 29.21(91.27) | -16.89(73.27) | -43.01(63.62) | -30.73(62.40) | 13.44(81.79) |
| **Blood sampling (t = 60 minute)** | | | | | | | | |
| **Lipid Class** | **Average lipid level in Hepatic vein (S.D.), ×10$^{02}$** | | | | **Average lipid level in Systemic vein (S.D. ), ×10$^{02}$** | | | |
| | **All** | **CTL** | **ALD** | **NAFLD** | **All** | **CTL** | **ALD** | **NAFLD** |
| **Cer** | 0.83(56.44) | 15.23(59.09) | -16.15(49.17) | 7.07(60.64) | 2.13(63.20) | 0.81(81.81) | -1.78(51.08) | 6.64(63.86) |
| **DG** | 5.81(58.82) | -5.15(54.65) | -13.03(53.52) | 30.69(60.89) | 1.06(48.22) | -14.93(52.90) | -11.61(46.04) | 23.55(40.74) |
| **FFA** | -49.10(45.54) | -50.79(35.16) | -73.59(41.93) | -25.11(44.54) | 17.67(47.75) | 9.11(40.71) | 15.24(56.21) | 25.63(45.41) |
| **HexCer** | 3.48(65.69) | 30.55(78.79) | 7.06(53.17) | -17.91(63.80) | -2.38(79.11) | 10.54(133.52) | 9.90(45.08) | -22.45(54.18) |
| **LacCer** | -2.84(79.92) | 36.80(81.56) | -29.08(70.04) | -4.76(81.80) | 4.06(78.71) | 41.02(93.51) | -9.29(67.84) | -8.12(74.90) |
| **LPC** | -9.05(80.53) | 39.53(27.12) | -11.73(91.10) | -38.93(82.31) | 7.56(74.81) | 55.37(30.55) | 1.93(77.21) | -19.06(81.42) |
| **LPE** | -4.23(64.23) | 13.51(30.69) | -4.87(62.59) | -15.47(81.20) | 6.21(59.09) | 15.60(43.48) | 16.43(67.12) | -9.58(60.38) |
| **PC** | -3.50(39.49) | 18.76(36.32) | -12.21(31.39) | -10.21(44.63) | 6.32(44.60) | 24.36(44.04) | 3.74(38.48) | -3.30(49.39) |
| **PE** | -1.32(46.29) | 4.83(43.87) | -8.98(59.66) | 1.73(34.33) | 5.24(53.98) | 13.18(51.96) | -0.25(71.65) | 5.08(36.50) |
| **PI** | -2.08(63.29) | -18.92(44.38) | 7.30(61.35) | 0.40(76.24) | 6.96(62.57) | -25.48(56.76) | 34.03(43.56) | 3.32(72.77) |
| **SHexCer** | 16.86(50.56) | 13.54(59.96) | 22.45(40.22) | 13.86(55.48) | 2.88(90.57) | -42.02(136.33) | 33.60(66.70) | 4.15(61.64) |
| **SM** | -4.21(75.58) | 45.92(80.42) | -51.65(55.41) | 6.64(65.75) | 5.61(71.74) | 48.74(79.68) | -35.11(52.26) | 14.85(65.39) |
| **TG** | 0.88(73.13) | -57.62(49.35) | -10.49(48.81) | 50.50(74.34) | 4.67(75.58) | -62.24(57.00) | 8.03(57.39) | 46.13(72.86) |
| **Blood sampling (t = 180 minute)** | | | | | | | | |
| **Lipid Class** | **Average lipid level in Hepatic vein (S.D.), ×10$^{02}$** | | | | **Average lipid level in Systemic vein (S.D. ), ×10$^{02}$** | | | |
| | **All** | **CTL** | **ALD** | **NAFLD** | **All** | **CTL** | **ALD** | **NAFLD** |
| **Cer** | 0.99(59.43) | 10.16(58.13) | -4.30(61.29) | -0.19(61.93) | -5.61(63.89) | -9.09(79.55) | -5.38(57.63) | -3.50(62.56) |
| **DG** | -2.19(46.55) | -10.05(40.89) | -13.10(42.16) | 13.23(52.32) | -13.23(49.54) | -29.98(50.81) | -13.28(43.19) | -2.00(54.18) |
| **FFA** | -65.71(52.59) | -57.75(27.70) | -72.13(46.80) | -65.03(69.95) | 4.02(48.03) | 8.53(53.55) | 1.24(56.45) | 3.60(37.88) |
| **HexCer** | 2.48(64.82) | 19.63(81.91) | 9.24(46.07) | -15.26(67.29) | -8.29(87.96) | -11.99(131.99) | 14.89(61.61) | -27.47(73.04) |
| **LacCer** | -4.83(85.03) | 47.98(76.17) | -23.67(67.99) | -22.45(94.46) | 6.65(73.63) | 47.57(65.97) | -12.04(65.55) | -3.19(79.24) |
| **LPC** | -21.35(83.34) | 4.57(26.94) | -22.78(111.62) | -37.30(77.85) | -5.14(72.63) | 22.04(20.99) | -4.92(85.80) | -23.46(79.45) |
| **LPE** | -4.75(75.47) | -5.85(48.89) | 11.20(92.67) | -18.91(74.02) | 3.28(50.95) | 6.50(32.76) | 14.50(55.24) | -9.35(56.74) |
| **PC** | -7.69(43.81) | 7.61(32.98) | -8.36(47.57) | -17.25(46.29) | 0.56(41.53) | 18.29(31.35) | -2.31(41.01) | -8.60(46.53) |
| **PE** | -12.13(53.65) | -0.10(52.26) | -15.53(71.11) | -16.97(34.96) | 3.07(51.46) | 5.57(43.44) | 7.20(71.49) | -2.45(34.31) |
| **PI** | -4.49(67.09) | -9.45(40.80) | 11.69(63.07) | -16.29(83.82) | -0.34(62.86) | -28.35(67.64) | 22.31(54.48) | -2.81(62.84) |
| **SHexCer** | 5.95(67.11) | -16.13(104.82) | 26.52(48.07) | 1.48(47.14) | -22.99(176.83) | -122.43(316.53) | 19.62(86.16) | 3.52(62.72) |
| **SM** | -8.72(77.42) | 37.99(75.12) | -49.44(73.05) | -1.85(66.47) | 0.26(68.56) | 42.20(68.10) | -36.49(49.02) | 6.60(69.97) |
| **TG** | 9.58(53.85) | -23.00(38.84) | 10.79(43.64) | 30.18(62.60) | 15.19(62.39) | -25.22(37.37) | 9.77(49.33) | 47.20(71.51) |

**Supplementary Table 3. Relative levels of 13 lipid classes in hepatic vein and systemic vein at 3 timepoints (blood sampling at t=0 min, t=60 min and t=180 min).**
Abbreviation. S.D., Standard deviation; CI, Confidence interval; CTL, Control healthy; ALD, Alcohol related liver disease; NAFLD, Non-alcohol related fatty liver disease
*Different to CTL at Bonferroni procedure $p<3.85\times10^{-03}$ level according to mixed model.
[#]Different between hepatic and systemic values at Bonferroni procedure $p<0.0039$ level according to mixed model.

| Fixed Effect | 1. Time | | 2. Time | | 3. Time*(Systemic vs. Hepatic) | |
|---|---|---|---|---|---|---|
| **Vein Site** | **Hepatic** | | **Systemic** | | **Systemic vs. Hepatic** | |
| **Variable** | **Est.Coef. (95% CI)** | **Slope p-value** | **Est.Coef. (95% CI)** | **Slope p-value** | **Slope p-value** | **Baseline p-value** |
| **FFA(18:1)** | -64.75 (-82.01~-47.50) | *$1.05\times10^{-10}$ | -60.57 (-80.83~-40.32) | *$7.27\times10^{-08}$ | $7.43\times10^{-01}$ | *$1.34\times10^{-08}$ |
| **FFA(18:2)** | -82.01 (-98.99~-65.03) | *$7.82\times10^{-15}$ | -65.29 (-80.37~-50.20) | *$6.57\times10^{-13}$ | $1.30\times10^{-01}$ | *$7.44\times10^{-18}$ |
| **FFA(22:6)** | -53.16 (-68.91~-37.42) | *$2.76\times10^{-09}$ | -39.91 (-50.80~-29.03) | *$2.25\times10^{-10}$ | $1.57\times10^{-01}$ | *$5.08\times10^{-12}$ |
| **FFA(20:4)** | -50.70 (-67.35~-34.05) | *$4.62\times10^{-08}$ | -46.60 (-58.99~-34.22) | *$9.63\times10^{-11}$ | $6.88\times10^{-01}$ | *$2.00\times10^{-18}$ |
| **TG(50:2)** | 23.13 (11.44~34.83) | *$1.78\times10^{-04}$ | 27.36 (16.30~38.43) | *$4.70\times10^{-06}$ | $5.87\times10^{-01}$ | $1.64\times10^{-01}$ |
| **TG(50:3)** | 28.22 (15.82~40.63) | *$2.13\times10^{-05}$ | 31.95 (17.10~46.80) | *$5.24\times10^{-05}$ | $6.91\times10^{-01}$ | $5.04\times10^{-02}$ |
| **TG(52:1)** | 22.98 (11.46~34.49) | *$1.58\times10^{-04}$ | 28.06 (18.73~37.40) | *$6.39\times10^{-08}$ | $4.91\times10^{-01}$ | $1.66\times10^{-01}$ |
| **LPC(18:2)** | -37.76 (-46.76~-28.77) | *$2.06\times10^{-12}$ | -35.81 (-43.07~-28.55) | *$3.17\times10^{-15}$ | $7.27\times10^{-01}$ | $1.25\times10^{-02}$ |
| **LPC(18:1)** | -21.87 (-29.73~-14.01) | *$3.99\times10^{-07}$ | -19.98 (-26.89~-13.07) | *$1.66\times10^{-07}$ | $7.05\times10^{-01}$ | $1.17\times10^{-01}$ |
| **TG(48:1)** | 28.87 (18.51~39.22) | *$3.86\times10^{-07}$ | 33.01 (23.87~42.15) | *$3.62\times10^{-10}$ | $5.35\times10^{-01}$ | $7.80\times10^{-02}$ |
| **TG(48:2)** | 35.26 (24.04~46.48) | *$2.03\times10^{-08}$ | 40.67 (30.51~50.83) | *$1.15\times10^{-11}$ | $4.53\times10^{-01}$ | $2.30\times10^{-02}$ |
| **LPC(20:4)** | -25.25 (-33.20~-17.30) | *$1.54\times10^{-08}$ | -26.33 (-33.85~-18.81) | *$9.43\times10^{-10}$ | $8.40\times10^{-01}$ | $4.67\times10^{-02}$ |
| **TG(46:1)** | 29.53 (16.98~42.09) | *$1.19\times10^{-05}$ | 34.61 (25.32~43.89) | *$1.29\times10^{-10}$ | $5.18\times10^{-01}$ | $5.74\times10^{-02}$ |
| **TG(46:2)** | 40.41 (22.81~58.01) | *$1.81\times10^{-05}$ | 43.92 (32.84~55.01) | *$1.65\times10^{-11}$ | $7.19\times10^{-01}$ | $2.14\times10^{-02}$ |
| **TG(46:0)** | 24.32 (14.03~34.60) | *$1.08\times10^{-05}$ | 30.50 (20.14~40.86) | *$1.08\times10^{-07}$ | $3.91\times10^{-01}$ | $8.45\times10^{-02}$ |
| **LPC(20:3/)** | -35.56 (-45.89~-25.23) | *$1.56\times10^{-09}$ | -43.95 (-53.57~-34.33) | *$7.84\times10^{-14}$ | $2.11\times10^{-01}$ | $1.39\times10^{-03}$ |
| **LPC(22:6)** | -36.93 (-47.75~-26.12) | *$1.98\times10^{-09}$ | -44.26 (-54.46~-34.05) | *$6.05\times10^{-13}$ | $3.07\times10^{-01}$ | $4.92\times10^{-04}$ |
| **TG(44:1)** | 36.11 (23.55~48.67) | *$1.90\times10^{-07}$ | 42.26 (31.40~53.12) | *$3.09\times10^{-11}$ | $4.37\times10^{-01}$ | $1.40\times10^{-02}$ |
| **TG(44:0)** | 26.43 (14.99~37.87) | *$1.63\times10^{-05}$ | 29.10 (17.95~40.26) | *$1.63\times10^{-06}$ | $7.27\times10^{-01}$ | $1.06\times10^{-01}$ |
| **PC(30:1)** | 14.03 (8.17~19.88) | *$8.50\times10^{-06}$ | 9.80 (1.59~18.02) | $2.00\times10^{-02}$ | $4.01\times10^{-01}$ | $7.26\times10^{-01}$ |
| **LPC(20:5)** | -23.67 (-31.19~-16.16) | *$1.92\times10^{-08}$ | -27.47 (-34.07~-20.86) | *$2.93\times10^{-12}$ | $4.28\times10^{-01}$ | $7.10\times10^{-02}$ |
| **LPC(/20:3)** | -21.52 (-30.76~-12.29) | *$1.39\times10^{-05}$ | -29.18 (-47.23~-11.14) | $1.88\times10^{-03}$ | $4.44\times10^{-01}$ | $8.69\times10^{-02}$ |
| **LPC(22:5/)** | -30.15 (-43.45~-16.85) | *$2.26\times10^{-05}$ | -47.00 (-61.41~-32.60) | *$7.37\times10^{-09}$ | $1.09\times10^{-01}$ | $1.44\times10^{-03}$ |
| **LPC(/22:5)** | -32.48 (-46.38~-18.57) | *$1.35\times10^{-05}$ | -41.74 (-56.00~-27.48) | *$1.24\times10^{-07}$ | $3.28\times10^{-01}$ | $1.48\times10^{-03}$ |

**Supplementary Table 4. Summary of mixed models investigation association between selected 24 lipids and 3 different fixed effects.** The 24 lipids shown in this table were selected as their hepatic vein levels associated with time at Bonferroni procedure $p<1.98\times10^{-04}$ level. The fixed effects shown in this table includes (1) Time from hepatic vein, (2) Time from systemic vein, and (3) Time*(Systemic vs. Hepatic). When fixed effect consists of a single variable (e.g., Time), estimate coefficient values (change in relative level of lipid every minute after alcohol intervention) and their p-values were calculated. When fixed effect is an interaction between variables (e.g., Time*(CTL vs. ALD)), slope p-values representing significant differences between changes in lipid levels between groups, and baseline p-values representing significant differences between lipid levels at baseline (time = 0min) were derived.
**Abbreviation**. FFA, Free fatty acid; TG, Triglyceride; LPC, Lysophosphatidylcholine; PC, Phosphatdiylcholine; Est.Coef., Estimate coefficient; CI, Confidence interval
*Passed Bonferroni procedure $p<1.98\times10^{-04}$.

| Fixed Effect | 1. Time in CTL | | 2. Time in ALD | | 3. Time in NAFLD | | 4. Time*(CTL vs. ALD) | | 5. Time*(CTL vs. NAFLD) | | 6. Time*(ALD vs. NAFLD) | |
|---|---|---|---|---|---|---|---|---|---|---|---|---|
| **Vein Site** | **Hepatic** | | **Hepatic** | | **Hepatic** | | **Hepatic** | | **Hepatic** | | **Hepatic** | |
| **Variable** | **Est.Coef. (95% CI)** | **Slope p-value** | **Est.Coef. (95% CI)** | **Slope p-value** | **Est.Coef. (95% CI)** | **Slope p-value** | **Slope p-value** | **Baseline p-value** | **Slope p-value** | **Baseline p-value** | **Slope p-value** | **Baseline p-value** |
| **FFA(18:1)** | -66.15 (-90.65~-41.65) | *$1.90\times10^{-05}$ | -71.91 (-100.66~-43.16) | *$2.14\times10^{-05}$ | -57.14 (-91.62~-22.66) | $2.03\times10^{-03}$ | $7.67\times10^{-01}$ | $2.58\times10^{-01}$ | $6.94\times10^{-01}$ | $2.75\times10^{-01}$ | $5.07\times10^{-01}$ | $3.66\times10^{-02}$ |
| **FFA(18:2)** | -77.66 (-103.13~-52.20) | *$4.02\times10^{-06}$ | -96.61 (-131.11~-62.11) | *$4.14\times10^{-06}$ | -71.27 (-98.75~-43.80) | *$1.08\times10^{-05}$ | $4.02\times10^{-01}$ | $5.34\times10^{-02}$ | $7.41\times10^{-01}$ | $3.33\times10^{-02}$ | $2.41\times10^{-01}$ | $3.76\times10^{-01}$ |
| **FFA(22:6)** | -36.12 (-69.81~-2.44) | $3.69\times10^{-02}$ | -60.13 (-87.73~-32.52) | *$1.27\times10^{-04}$ | -58.03 (-83.25~-32.80) | *$5.76\times10^{-05}$ | $2.58\times10^{-01}$ | $3.72\times10^{-01}$ | $2.79\times10^{-01}$ | $3.64\times10^{-01}$ | $9.09\times10^{-01}$ | $7.23\times10^{-01}$ |
| **FFA(20:4)** | -44.59 (-74.38~-14.81) | $5.47\times10^{-03}$ | -71.97 (-102.17~-41.78) | *$4.09\times10^{-05}$ | -34.91 (-62.49~-7.33) | $1.49\times10^{-02}$ | $2.03\times10^{-01}$ | $1.70\times10^{-01}$ | $6.34\times10^{-01}$ | $1.44\times10^{-01}$ | $6.81\times10^{-02}$ | $9.63\times10^{-01}$ |
| **TG(50:2)** | 49.33 (25.00~73.66) | $4.39\times10^{-04}$ | 22.15 (7.54~36.76) | $4.38\times10^{-03}$ | 6.59 (-14.84~28.01) | $5.34\times10^{-01}$ | $4.10\times10^{-02}$ | *$1.44\times10^{-07}$ | $1.01\times10^{-02}$ | *$2.62\times10^{-05}$ | $2.31\times10^{-01}$ | $4.37\times10^{-01}$ |
| **TG(50:3)** | 48.65 (23.60~73.70) | $6.61\times10^{-04}$ | 36.45 (21.54~51.36) | *$2.91\times10^{-05}$ | 6.92 (-16.59~30.43) | $5.52\times10^{-01}$ | $3.62\times10^{-01}$ | *$2.19\times10^{-05}$ | $1.87\times10^{-02}$ | $1.70\times10^{-03}$ | $3.70\times10^{-02}$ | $6.11\times10^{-01}$ |
| **TG(52:1)** | 42.04 (22.53~61.55) | $2.40\times10^{-04}$ | 28.61 (16.49~40.74) | *$4.67\times10^{-05}$ | 5.01 (-19.37~29.39) | $6.77\times10^{-01}$ | $2.09\times10^{-01}$ | *$6.16\times10^{-07}$ | $2.94\times10^{-02}$ | *$8.09\times10^{-06}$ | $8.82\times10^{-02}$ | $7.23\times10^{-01}$ |
| **LPC(18:2)** | -43.13 (-54.70~-31.56) | *$2.43\times10^{-07}$ | -49.19 (-68.44~-29.93) | *$1.60\times10^{-05}$ | -23.53 (-36.32~-10.73) | $7.63\times10^{-04}$ | $6.18\times10^{-01}$ | $8.59\times10^{-03}$ | $3.26\times10^{-02}$ | $1.19\times10^{-03}$ | $2.50\times10^{-02}$ | $3.32\times10^{-01}$ |
| **LPC(18:1)** | -32.61 (-43.83~-21.39) | *$7.51\times10^{-06}$ | -29.97 (-45.23~-14.71) | $4.09\times10^{-04}$ | -7.15 (-19.17~4.88) | $2.34\times10^{-01}$ | $7.91\times10^{-01}$ | $7.99\times10^{-02}$ | $4.06\times10^{-03}$ | $6.98\times10^{-02}$ | $1.86\times10^{-02}$ | $1.56\times10^{-01}$ |
| **TG(48:1)** | 58.89 (37.99~79.79) | *$1.11\times10^{-05}$ | 29.21 (16.87~41.55) | *$4.47\times10^{-05}$ | 8.53 (-9.38~26.44) | $3.38\times10^{-01}$ | $9.85\times10^{-03}$ | *$1.30\times10^{-07}$ | $4.95\times10^{-04}$ | *$8.01\times10^{-06}$ | $6.00\times10^{-02}$ | $8.99\times10^{-01}$ |
| **TG(48:2)** | 64.32 (40.34~88.29) | *$2.05\times10^{-05}$ | 39.07 (25.41~52.73) | *$3.00\times10^{-06}$ | 12.33 (-6.55~31.22) | $1.92\times10^{-01}$ | $4.87\times10^{-02}$ | *$4.53\times10^{-07}$ | $8.94\times10^{-04}$ | *$6.03\times10^{-05}$ | $2.40\times10^{-02}$ | $8.71\times10^{-01}$ |
| **LPC(20:4)** | -36.35 (-48.13~-24.57) | *$3.43\times10^{-06}$ | -32.75 (-47.54~-17.96) | *$1.04\times10^{-04}$ | -10.84 (-23.66~1.97) | $9.42\times10^{-02}$ | $7.14\times10^{-01}$ | $7.16\times10^{-02}$ | $6.46\times10^{-03}$ | $5.75\times10^{-02}$ | $2.51\times10^{-02}$ | $7.77\times10^{-01}$ |
| **TG(46:1)** | 46.20 (17.04~75.36) | $3.63\times10^{-03}$ | 40.26 (18.55~61.96) | $7.39\times10^{-04}$ | 8.41 (-8.33~25.16) | $3.13\times10^{-01}$ | $7.31\times10^{-01}$ | *$2.94\times10^{-05}$ | $1.61\times10^{-02}$ | *$1.56\times10^{-04}$ | $1.98\times10^{-02}$ | $4.11\times10^{-01}$ |
| **TG(46:2)** | 67.81 (44.08~91.54) | *$9.33\times10^{-06}$ | 66.77 (35.38~98.15) | *$1.68\times10^{-04}$ | -2.46 (-29.66~24.73) | $8.54\times10^{-01}$ | $9.60\times10^{-01}$ | *$5.07\times10^{-05}$ | $4.81\times10^{-04}$ | $4.44\times10^{-04}$ | $1.15\times10^{-03}$ | $2.14\times10^{-02}$ |
| **TG(46:0)** | 52.40 (34.45~70.35) | *$7.11\times10^{-06}$ | 27.16 (14.75~39.57) | *$1.20\times10^{-04}$ | 2.94 (-16.03~21.91) | $7.54\times10^{-01}$ | $1.69\times10^{-02}$ | *$8.89\times10^{-08}$ | $5.51\times10^{-04}$ | *$3.08\times10^{-05}$ | $3.55\times10^{-02}$ | $7.21\times10^{-01}$ |
| **LPC(20:3/)** | -52.16 (-67.48~-36.85) | *$8.90\times10^{-07}$ | -43.26 (-63.30~-23.21) | *$1.42\times10^{-04}$ | -17.31 (-33.10~-1.51) | $3.28\times10^{-02}$ | $5.01\times10^{-01}$ | $9.86\times10^{-02}$ | $3.12\times10^{-03}$ | $6.28\times10^{-02}$ | $4.04\times10^{-02}$ | $3.96\times10^{-01}$ |
| **LPC(22:6)** | -45.83 (-57.40~-34.26) | *$9.80\times10^{-08}$ | -46.49 (-70.91~-22.06) | $5.70\times10^{-04}$ | -22.08 (-37.21~-6.95) | $5.72\times10^{-03}$ | $9.65\times10^{-01}$ | $2.53\times10^{-03}$ | $2.33\times10^{-02}$ | $1.29\times10^{-03}$ | $8.27\times10^{-02}$ | $9.11\times10^{-01}$ |
| **TG(44:1)** | 73.08 (54.47~91.68) | *$1.12\times10^{-07}$ | 43.68 (23.47~63.89) | *$1.39\times10^{-04}$ | 4.40 (-14.77~23.57) | $6.42\times10^{-01}$ | $3.96\times10^{-02}$ | *$2.17\times10^{-07}$ | *$6.76\times10^{-06}$ | *$4.55\times10^{-05}$ | $5.47\times10^{-03}$ | $1.70\times10^{-01}$ |
| **TG(44:0)** | 51.92 (34.38~69.46) | *$5.95\times10^{-06}$ | 37.92 (20.58~55.25) | *$1.20\times10^{-04}$ | -1.28 (-20.64~18.07) | $8.93\times10^{-01}$ | $2.59\times10^{-01}$ | *$3.99\times10^{-06}$ | $2.57\times10^{-04}$ | $7.98\times10^{-04}$ | $3.26\times10^{-03}$ | $1.67\times10^{-01}$ |
| **PC(30:1)** | 27.99 (14.38~41.60) | $3.82\times10^{-04}$ | 10.25 (0.96~19.54) | $3.18\times10^{-02}$ | 8.24 (-0.26~16.73) | $5.69\times10^{-02}$ | $2.52\times10^{-02}$ | $7.32\times10^{-03}$ | $9.82\times10^{-03}$ | $2.81\times10^{-03}$ | $7.44\times10^{-01}$ | $1.34\times10^{-01}$ |
| **LPC(20:5)** | -31.78 (-40.99~-22.58) | *$7.36\times10^{-07}$ | -32.63 (-45.05~-20.22) | *$1.06\times10^{-05}$ | -9.90 (-23.99~4.19) | $1.61\times10^{-01}$ | $9.17\times10^{-01}$ | $1.41\times10^{-01}$ | $2.13\times10^{-02}$ | $1.94\times10^{-01}$ | $1.68\times10^{-02}$ | $2.71\times10^{-01}$ |
| **LPC(/20:3)** | -34.98 (-45.82~-24.14) | *$1.89\times10^{-06}$ | -33.77 (-51.95~-15.58) | $7.32\times10^{-04}$ | -1.13 (-14.86~12.61) | $8.68\times10^{-01}$ | $9.16\times10^{-01}$ | $5.23\times10^{-02}$ | $6.37\times10^{-04}$ | $2.38\times10^{-02}$ | $4.51\times10^{-03}$ | $6.74\times10^{-01}$ |
| **LPC(22:5/)** | -52.99 (-87.57~-18.41) | $4.64\times10^{-03}$ | -36.05 (-54.92~-17.17) | $5.49\times10^{-04}$ | -9.41 (-28.52~9.70) | $3.22\times10^{-01}$ | $3.43\times10^{-01}$ | $1.61\times10^{-01}$ | $1.72\times10^{-02}$ | $1.65\times10^{-01}$ | $4.73\times10^{-02}$ | $1.47\times10^{-01}$ |
| **LPC(/22:5)** | -41.81 (-55.95~-27.67) | *$6.04\times10^{-06}$ | -39.76 (-72.56~-6.96) | $1.94\times10^{-02}$ | -19.46 (-38.32~-0.59) | $4.36\times10^{-02}$ | $9.18\times10^{-01}$ | $2.57\times10^{-01}$ | $8.22\times10^{-02}$ | $2.17\times10^{-01}$ | $2.68\times10^{-01}$ | $2.25\times10^{-01}$ |

**Supplementary Table 5. Summary of mixed models investigation association between selected 24 lipids and 6 different fixed effects.** The 24 lipids in this table are same as those found in Supp. Table 2. The fixed effects shown in this table includes (1) Time from CTL hepatic, (2) Time from ALD hepatic, and (3) Time from NAFLD hepatic, (4) Time*(CTL vs. ALD) from hepatic, (5) Time*(CTL vs. NAFLD) from hepatic, and (6) Time*( ALD vs. NAFLD) from hepatic. When fixed effect consists of a single variable (e.g., Time), estimate coefficient (change in relative level of lipid every minute after alcohol intervention) and their p-values were calculated. When fixed effect is an interaction between variables (e.g., Time*(CTL vs. ALD)), slope p-values representing significant differences between changes in lipid levels between groups, and baseline p-values representing significant differences between lipid levels at baseline (time = 0min) were derived.

**Abbreviation**. FFA, Free fatty acid; TG, Triglyceride; LPC, Lysophosphatidylcholine; PC, Phosphatdiylcholine; Est.Coef., Estimate coefficient; CI, Confidence interval; CTL, Control healthy; ALD, Alcohol related liver disease; NAFLD, Non-alcohol related fatty liver disease*Passed Bonferroni procedure $p<1.98\times10^{-04}$.
